# Unveiling the Burden of Drug-Induced Impulsivity: A Network Analysis of the FDA Adverse Event Reporting System

**DOI:** 10.1101/2023.11.17.23298635

**Authors:** Michele Fusaroli, Stefano Polizzi, Luca Menestrina, Valentina Giunchi, Luca Pellegrini, Emanuel Raschi, Daniel Weintraub, Maurizio Recanatini, Gastone Castellani, Fabrizio De Ponti, Elisabetta Poluzzi

## Abstract

**Introduction:** Impulsivity induced by dopaminergic agents, like pramipexole and aripiprazole, can lead to behavioral addictions impacting social functioning and quality of life of patients and families (e.g., resulting in unemployment, marital problems, anxiety). These secondary effects, interconnected in networks of signs and symptoms, are usually overlooked by clinical trials, not reported in package inserts, and neglected in clinical practice.

**Objective:** This study explores the syndromic burden of impulsivity induced by pramipexole and aripiprazole, pinpointing key symptoms for targeted mitigation.

**Methods:** An event-event Information Component (IC) on the FDA Adverse Event Reporting System (January 2004 – March 2022) identified the syndrome of events disproportionally co-reported with impulsivity, separately for pramipexole and aripiprazole. A greedy-modularity clustering on composite network analyses (PPMI, Ising, Φ) identified subsyndromes. Bayesian network modeling highlighted possible precipitating events.

**Results:** Suspected drug-induced impulsivity was documented in 7.49% pramipexole and 4.50% aripiprazole recipients. The highest IC concerned obsessive-compulsive disorder (reporting rate = 26.77%; IC median = 3.47, 95%CI = 3.33-3.57) and emotional distress (21.35%; 3.42, 3.26-3.54) for pramipexole, bankruptcy (10.58%; 4.43, 4.26-4.55) and divorce (7.59%; 4.38, 4.19-4.53) for aripiprazole. The network analysis identified delusional jealousy and dopamine dysregulation subsyndromes for pramipexole, obesity-hypoventilation and social issues for aripiprazole. The Bayesian network highlighted anxiety and economic problems as potentially precipitating events.

**Conclusion:** The under-explored consequences of drug-induced impulsivity significantly burden patients and families. Network analyses, exploring syndromic reactions and potential precipitating events, complement traditional techniques and clinical judgment. Characterizing the secondary impact of reactions will support informed patient-centered decision-making.

**Highlights:**

- Drug-induced impulsivity significantly impacts patients’ lives. Network analyses help characterize reactions as syndromes.
- We explore the impulsivity syndrome and subsyndromes resulting from pramipexole and aripiprazole.
- The manifestation of drug-induced impulsivity was different for the two drugs. Anxiety and economic problems bridge between other symptoms and could be important therapeutical targets.

## 1. Introduction

Adverse drug reactions (ADRs) significantly impact patients’ well-being [1], extending from organic to psychological and social issues [2,3]. For instance, immunodeficiency perturbs social activities, dysphonia hinders the ability to function in positions requiring extensive vocal communication such as teaching or other public-facing roles, and sexual dysfunction can affect relationships and even personal identity, with further cascading effects on psychological and physical well-being. Despite their profound effects and complex networks of interactions, ADRs are often inadequately recognized, resulting in compromised patient-doctor relationships [4], prolonged hospitalization [5], and a pervasive decline in quality of life (QoL) [6]. Evidence obtained from patient-reported outcomes – crucial for QoL assessment and patient-centered care [7] – is equally disregarded and often relegated to the margins in prescribing information or package inserts [8].

Drug-induced impulsivity, classified as “impulse control disorders induced by other specified psychoactive substance (6C4E.73)” in the International Classification of Diseases (ICD-11) category of disorders due to substance use, represents a distressing group of conditions marked by a loss of behavioral control. This pathological disinhibition can yield behaviors as pervasive as pathologic gambling, hypersexuality, compulsive shopping, and hyperphagia–the so-called “four knights of Impulse Control Disorder”, all with important further repercussions on social relationships, physical and psychological wellbeing [9]. Other behaviors like stealing, hair pulling, and compulsive hoarding, while less acknowledged, can also occur, and further diversify potential manifestations of drug-induced impulsivity [10,11].

The first reports of drug-induced impulsivity were linked to dopamine receptor agonists like pramipexole, ropinirole, rotigotine, licensed for treating Parkinson’s disease (PD) [12] and restless legs syndrome (RLS) [13,14]. More recently, the role of partial dopamine agonists like aripiprazole, brexpiprazole, cariprazine, licensed for treating psychosis and mood disorders, has also emerged as cause of drug-induced impulsivity [15]. Impulsivity as induced by PD treatment has a complex trajectory and may have a different impact on quality of life depending on underlying susceptibilities and on the occurrence of exacerbating events. The treatment might initially induce a “honeymoon period”, in which patients experience heightened motivational drive and often engage in new satisfying hobbies [16]. However, impulsivity can eventually lead to pervasive behaviors and become pathologic. Even when concealed in subclinical forms [17], drug-induced impulsivity holds the potential to significantly erode patients’ QoL [18]. This erosion, appraised through metrics like the PDQ-39 scale [19], encompasses diverse neuropsychiatric and somatic domains including mobility, daily activities, stigma, social support, communication [20], urinary and sexual function, sleep, attention, and cardiovascular symptoms [21]. The impact extends beyond patients as it affects caregivers, who grapple with their own set of health issues, depression, and social impediments due to their duties of care [22].

Nevertheless, conventional evaluations focus on simple drug – physical adverse reaction associations, and often ignore neuropsychiatric symptoms, altered behavior patterns, financial hardships, and legal entanglements as emerging from drug-induced impulsivity [23], thus failing to capture the complex network of consequences of its manifestation. This underscores the crucial need for an integrative approach that consider the perspectives of both patients and caregivers and acknowledges the complex interconnections between symptoms [24], including identifying subsyndromes and exacerbating events.

In order to better characterize such networks, we can rely on databases collecting individual case safety reports of suspected ADRs from patients and healthcare professionals. In particular, in this manuscript we will rely on the US FDA Adverse Event Reporting System (FAERS), as it is global and public [25]. Crucially, the patient-produced reports offer impactful insights into the experiences and impacts of ADRs on QoL, beyond those provided by healthcare professionals [26–30].

Network analyses provide the means to explore ADRs as complex system consisting of multiple interacting entities. Specifically, they enable the analysis and visualization of ADRs as interwoven symptoms and signs [31]: a composite syndrome encompassing psychosocial implications, clustered into subsyndromes. This approach overcomes the drawbacks of viewing drug-induced impulsivity as an isolated incident. Beyond more descriptive approaches such as those built on measures of association (Ising [32], Φ [33], and PPMI [34]), network approaches can also explore possible causal connections between symptoms, allowing to identify potential exacerbating events [35,36].

In the current manuscript we pursue three goals. First, we want to fully acknowledge the complexity of drug-induced impulsivity as reported not only by clinical researchers, but also by clinicians and patients. We go beyond a pairwise drug to adverse reaction approach and more fully embrace the idea of a syndrome of variously interconnected aspects, from behavioral addictions to their organic and psychosocial sequelae. By examining the network in which symptoms interact and affect each other, we aim to gain a deeper understanding of the impact of drug-induced impulsivity on patients’ lives.

Second, we want to assess the possibility of distinct sub-syndromes within the broader spectrum of drug-induced impulsivity. For example, when impulsivity manifests as hyperphagia it may have more organic sequelae related to weight increased compared to when it manifests as pathological gambling.

Third, we assess the presence of central symptoms, potentially crucially exacerbating the syndrome of drug-induced impulsivity. By pinpointing these key symptoms, we aim to pave the way for targeted interventions that alleviate the adverse effects on patients’ lives.

In pursuing these goals, our study focuses on recipients of pramipexole and aripiprazole. Pramipexole is a dopamine agonist used in neurological conditions, therefore prescribed to older individual with stronger social support and a lower baseline motivational drive. Aripiprazole is a dopamine partial agonist employed in psychiatric disorders, therefore prescribed to younger and more stigmatized individuals, with a higher baseline motivational drive. Using these two examples will allow us to capture commonalities in drug-induced impulsivity as well as potentially different mechanisms/syndromes that manifest only in a specific indication/patient type. A better understanding of the impact of these ADRs on quality of life could contribute to informed decision-making for patients and caregivers, laying the foundation for interventions capable of alleviating the toll exacted by impulsivity.

## 2. Methods

### 2.0 Study Design

The study design is presented in **Figure 1**. We first downloaded and pre-processed FAERS reports from January 1^st^, 2004, to March 31^st^, 2022 (Step 1) and selected cases recording impulsivity among the two separate populations of aripiprazole and pramipexole recipients (Step 2). In order to characterize the impulsivity syndrome, we identified the set of events disproportionally co-reported with drug-induced impulsivity, rather than with other suspected reactions of the same drug, using an event-event disproportionality analysis (Step 3). To explore potential sub-syndromes, we relied on three parallel commonly used descriptive network analyses and a greedy-modularity algorithm, able to identify more cohesive clusters of co-reported events (Step 4). Finally, to explore possible causal relations within these networks and the secondary impact of drug-induced impulsivity, we relied on a Bayesian network approach, able to estimate conditional probabilities of chained events (Step 5). These operations were performed in a standardized and reproducible fashion, made possible by the DiAna R package: an open access toolkit for disproportionality analyses and other pharmacovigilance investigations in the FAERS [37].

**Figure 1.**
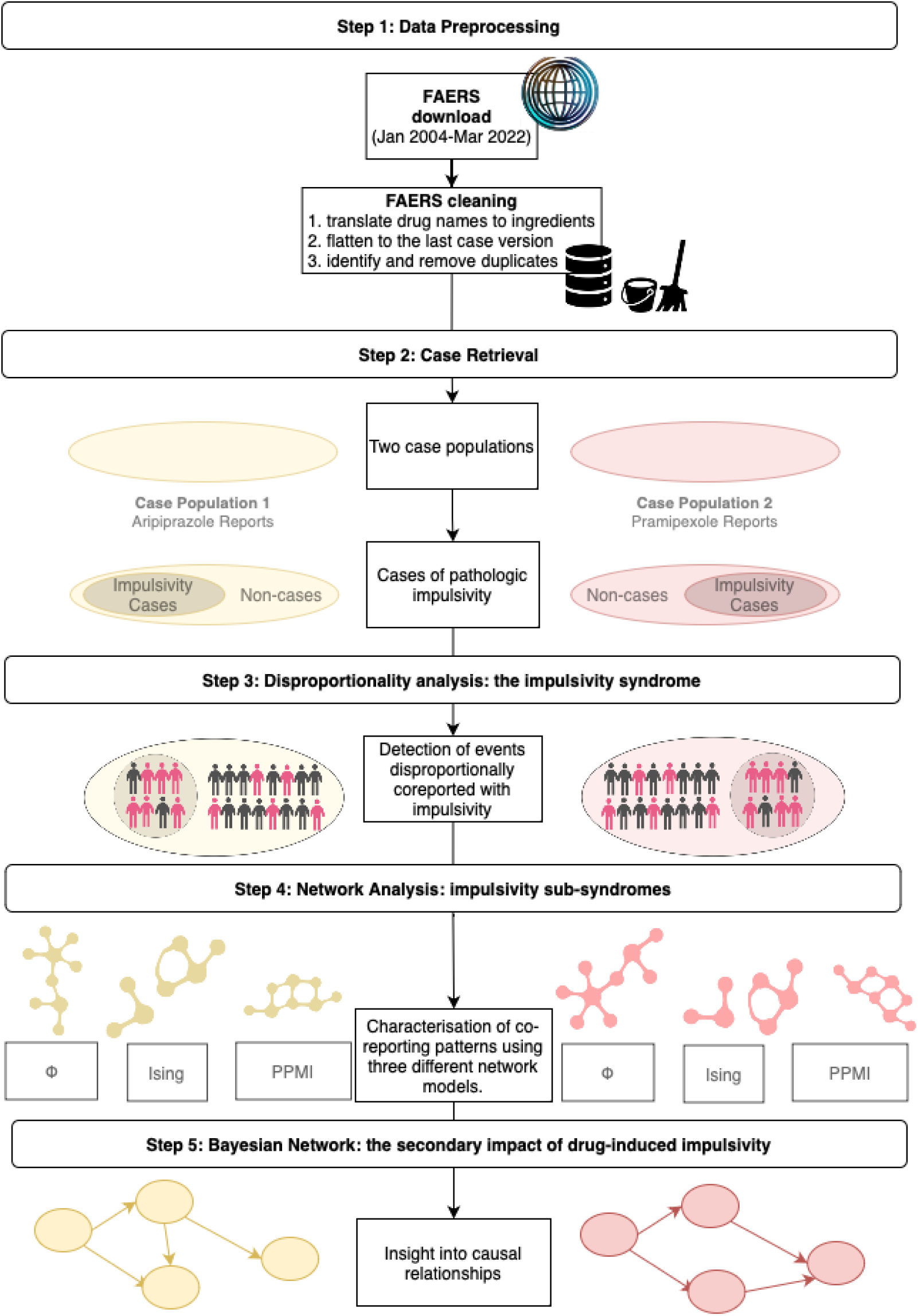
**Pipeline of the study**, showing step-by-step the study design.

### 2.1 Step 1 – Data Preprocessing

We downloaded FAERS quarterly data in ASCII format (January 1^st^, 2004, to March 31^st^, 2022) [25]. Adverse events were coded according to the Medical Dictionary for Regulatory Activities (MedDRA®, version 25.0) preferred terms (PTs) [38], while drugs were standardized according to their active ingredients [39]. The latest report version was retained, and rule-based deduplication was applied to reduce redundancy (cfr. https://github.com/fusarolimichele/DiAna).

### 2.2 Step 2 – Case Retrieval

We retrieved aripiprazole and pramipexole recipients separately and identified cases as reports recording impulsivity among the suspected reactions. To identify impulsivity we used a MedDRA PT list specifically curated for investigating drug-induced impulsivity within the FAERS database [10], encompassing heterogenous manifestations including gambling, hypersexuality, compulsive shopping, hyperphagia, gaming, setting fires, stealing, hoarding, excessive exercise, overwork, compulsive wandering, body-focused repetitive behaviors, stereotypy, and impulsivity (see **Table S1**). MedDRA PTs used for reporting suspected ADRs do not align directly with terms in other frameworks like the Diagnostic and Statistical Manual of mental disorders (DSM-5-TR) and ICD-11 (e.g., referring to “kleptomania” as an idiopathic condition).

To explore potential risk factors for impulsivity, demographic characteristics, outcomes, and reporter contributions (e.g., healthcare practitioners, patients, lawyers) were compared between cases and non-cases within each population, using the Chi-square test for categorical and Mann-Whitney test for continuous variables. To address multiple testing, we applied the Holm-Bonferroni correction with a significance level of 0.05.

### 2.3 Step 3 – Disproportionality Analysis: The drug-induced impulsivity syndrome

We conducted an event-event disproportionality analysis to identify events frequently co-reported with drug-induced impulsivity, separately for aripiprazole and pramipexole recipients (see **Table 1**). Disproportionate reporting was assessed using the Information Component (IC) [40], also known as pointwise mutual information (PMI) in information theory [41,42]. The IC compares the actual co-reporting of two events *x* (i.e., drug-induced impulsivity) and *y* (i.e., any specific event) with their expected co-reporting if their probability were independent (*p*(*y*, *x*) > *p*(*x*) * *p*(*y*)) [41]. To mitigate the risk of false positives for infrequent events [43], a shrinkage/smoothing approach was applied by adding k=0.5 to both the numerator and denominator. Significance was determined using *IC*_O25_ > 0.

**Table 1.** 2-way contingency table. The table shows the different instances that can be observed when considering pathologic impulsivity and a specific event. Legend: E = event; I = impulsivity; 1 = presence; 0 = lack; N = total

|  | Event (y) | Other events | Sum |
| --- | --- | --- | --- |
| **Impulsivity (x)** | $n_{I_1 E_1}$ | $n_{I_1 E_0}$ | $n_{I_1}$ |
| **Other event** | $n_{I_0 E_1}$ | $n_{I_0 E_0}$ | $n_{I_0}$ |
| **Sum** | $n_{E_1}$ | $n_{E_0}$ | **N** |

### 2.4 Step 4 – Network Analysis: sub-syndromes

Building on insights from prior studies on drugs [44] and events [31,45,46] co-occurrence, our network analysis aimed to unveil, within the drug-induced impulsivity syndrome, sub-syndromes of cohesively clustered events. Together with the Ising estimation [32,47], already implemented in pharmacovigilance because of its ability to obtain a sparser and easily interpretable network (because of fewer links and more definite clusters) [31], we implemented two other more connected network estimations in order to identify broader co-reporting patterns (PPMI [34], Ising, *ϕ* [33]). See **Table 2** for the features of the three networks. We did not consider negative links (i.e., potential mutually exclusive events).

**Table 2.** Network estimations. The table shows the features of the network estimations implemented.

| Ising | PPMI<br>(positive pointwise mutual information) | PHI |
| --- | --- | --- |
| The Ising model computed partial logistic regression coefficients ($\beta$) considering the impact of all other events. Positive coefficients indicated a tendency for two events to be reported together. A LASSO method pruned out weak links, eliminating spurious associations but potentially sacrificing weaker genuine relationships, especially triangular type interactions. | Focuses on cases where events were reported together ($n_{11}$), applying additive smoothing ($k=1$, $d=N^\circ$ events). Bootstrap and Bonferroni adjustments assessed statistical significance, with a 0.01 threshold for higher specificity. Gives more weight to associations with infrequent events. | The $\phi$ coefficient, akin to the traditional correlation coefficient, approaches one when two events are frequently reported together and converges to zero if they are either independent or mutually exclusive. The Bonferroni adjustment was applied with a significance threshold of 0.01. The p-value was computed using a $\chi^2$ probability distribution with one degree of freedom. |
| $Ising_{x,y} = \max\left(\frac{1}{2}\beta_{xy}\beta_{yx}, 0\right)$ | $PPMI_{x,y} = \max\left(\log_2 \frac{(n_{11} + k) \times (N + kd)}{(n_{1+} + k) \times (n_{+1} + k)}, 0\right)$ | $\phi_{x,y} = \max\left(\frac{n_{11}n_{00} - n_{10}n_{01}}{\sqrt{n_{1+}n_{0+}n_{+1}n_{+0}}}, 0\right)$ |

Separately for the two population of aripiprazole and pramipexole recipients, the three networks shared identical nodes by definition but different links were inferred due to the different properties of the algorithms. We used modularity maximization [48] and the greedy modularity algorithm [49] to detect clusters of co-reported signs and symptoms. Between networks we compared degree of link overlap (Jaccard similarity index [50]), goodness of partitioning (clustering modularity); cluster agreement (Purity index [51–53]), link density (ratio of actual links to possible links), and interconnectedness among neighbors (small worldness [54]).

### 2.5 Step 5 – Bayesian Network: The secondary impact of drug-induced impulsivity

While other methods look only at symmetrical associations, we expect that some events may cause other events. To exploratively identify these potential causal dependencies following drug-induced impulsivity, we estimated the conditional probabilities of chained events [35,36]. The resulting Bayesian network is both directed, offering insights into plausible causal relationships, and acyclic (no chain of arrows loops back to itself). The network was derived through 1000 bootstraps, optimizing the BIC score with the Hill-Climbing algorithm. We computed the average network retaining links exceeding a threshold computed via *L*_1_ minimization. Evaluation focused on nodes with the highest out-degree centrality and the main manifestations of drug-induced impulsivity.

## 3. Results

### 3.1 Case Retrieval

We retrieved 12,030,756 distinct reports: 27,601 pramipexole recipients and 80,238 aripiprazole recipients. Suspected drug-induced impulsivity was documented in 7.49% of pramipexole recipients (n=2,066: mainly gambling disorder–n=1,345; 4.87%–, hypersexuality–612; 2.22%–, impulsivity–453; 1.64%–, compulsive shopping–384; 1.39%–, and hyperphagia–334; 1.21%–) and in 4.50% aripiprazole recipients (n=3,609: mainly gambling disorder–n=2,067; 2.58%–, hypersexuality–1,077; 1.34%–, compulsive shopping– 1,029; 1.28%–, hyperphagia–868; 1.08%–, and impulsivity–730; 0.91%–) (see **Table S2**).

Among pramipexole recipients, drug-induced impulsivity was more frequently reported in males (57.42% vs. 36.99%, p<0.001) and younger patients (56 vs. 67, p<0.001), often recording non-serious outcomes (i.e., no death, disability, or hospitalization; 44.87% vs. 33.58%, p<0.001) and Parkinson’s Disease (PD) as indication (see **Figure 2** and **Table S3**). Similarly, among aripiprazole recipients, drug-induced impulsivity was more common in males (48.59% vs. 40.72%, p<0.001); peculiarly, hospitalization was more common (33.39% vs. 23.39%, p<0.001), and an important portion of reports was submitted by lawyers (34.08% vs. 1.10%, p<0.001) (see **Table S4**).

**Figure 2.**
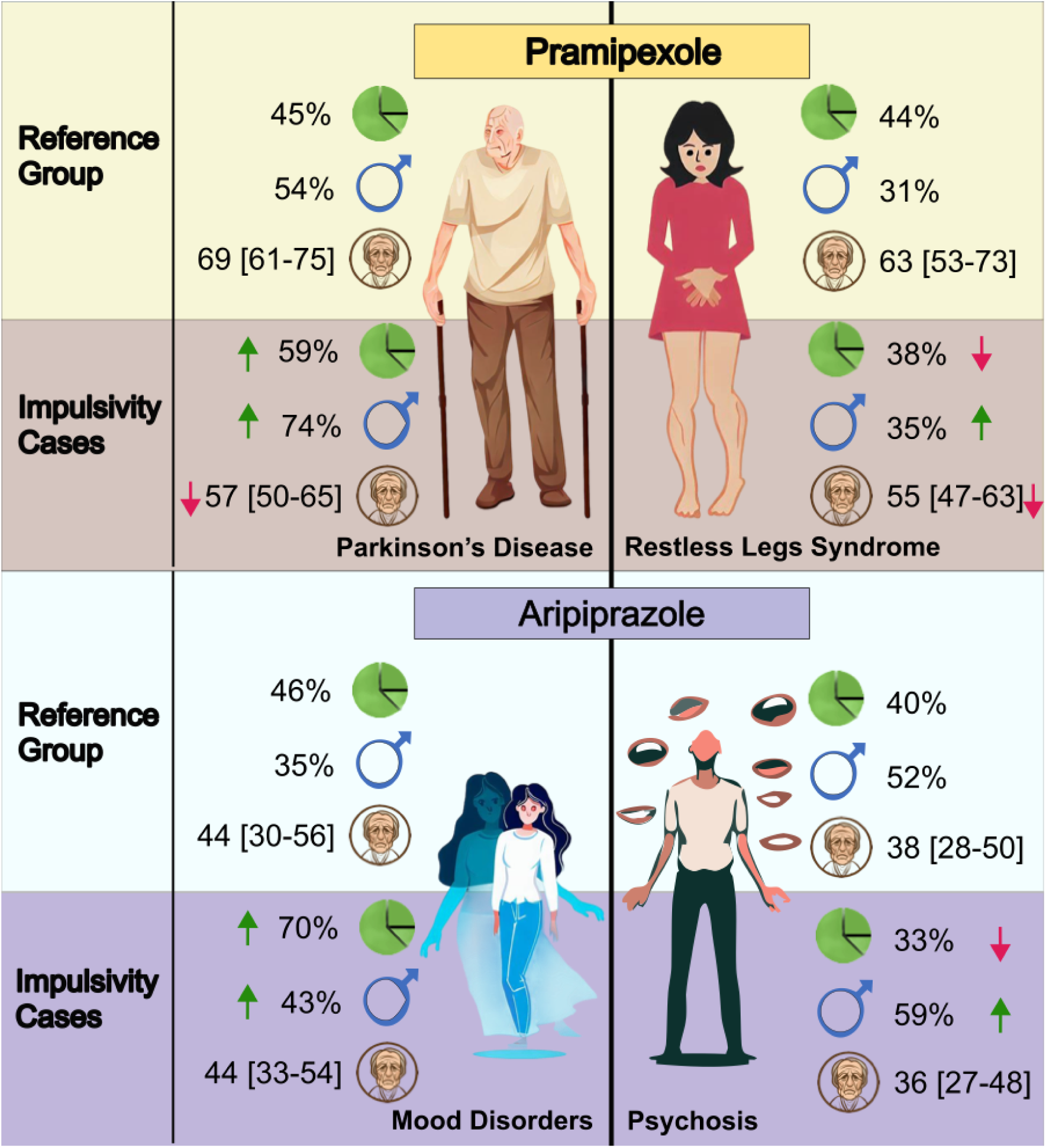
Characteristics of the investigated populations. The figure presents information about two populations extracted from the deduplicated FAERS database - one consisting of reports related to pramipexole and the other consisting of reports related to aripiprazole. Within these populations, cases of pathologic impulsivity were identified. The figure compares drug-induced impulsivity cases and the reference group (other reports recording the drug), considering the indication for use. Only the two most prevalent indications were taken into account. For each drug and indication, the caption describes the percentage of reports with the specified indication, the percent of reports involving males, and the median and interquartile range of ages. In the drug-induced impulsivity cases sections, green and red arrows indicate variables that are respectively higher or lower than expected based on the reference group.

### 3.2 Disproportionality Analysis: the drug-induced impulsivity syndrome

A total of 56 events were disproportionally reported with pramipexole-related impulsivity. The highest IC was found for obsessive-compulsive disorder (OCD, reporting rate = 26.77%; IC median = 3.47, 95%CI = 3.33-3.57), emotional distress (21.35%; 3.42, 3.26-3.54), marital problem (1.11%; 3.30, 2.61-3.79), dependence (2.37%; 3.26, 2.79-3.6), economic problems (6.05%; 3.15, 2.85-3.36), compulsions (1.74%; 3.05, 2.49-3.44), fear (4.65%; 2.95, 2.61-3.19), eating disorder (2.47%; 2.95, 2.49-3.28), personality change (2.66%; 2.93, 2.49-3.26), and suicide attempt (5.28%; 2.74, 2.43-2.97).

A total of 107 events were disproportionally reported with aripiprazole-related impulsivity. The highest IC was found for bankruptcy (10.58%; 4.43, 4.26-4.55), divorce (7.59%; 4.38, 4.19-4.53), homeless (6.93%; 4.37, 4.16-4.52), shoplifting (5.02%; 4.37, 4.12-4.54), neuropsychiatric symptoms (4.74%; 4.35, 4.1-4.53), loss of employment (12.64%; 4.33, 4.18-4.44), theft (5.79%; 4.32, 4.09-4.48), economic problems (37.85%; 4.28, 4.19-4.34), sexually transmitted disease (3.05%; 4.24, 3.93-4.47), and OCD (33.19%; 4.16, 4.07-4.23) (see **Figure 3 and Table S5 and S6**).

**Figure 3.**
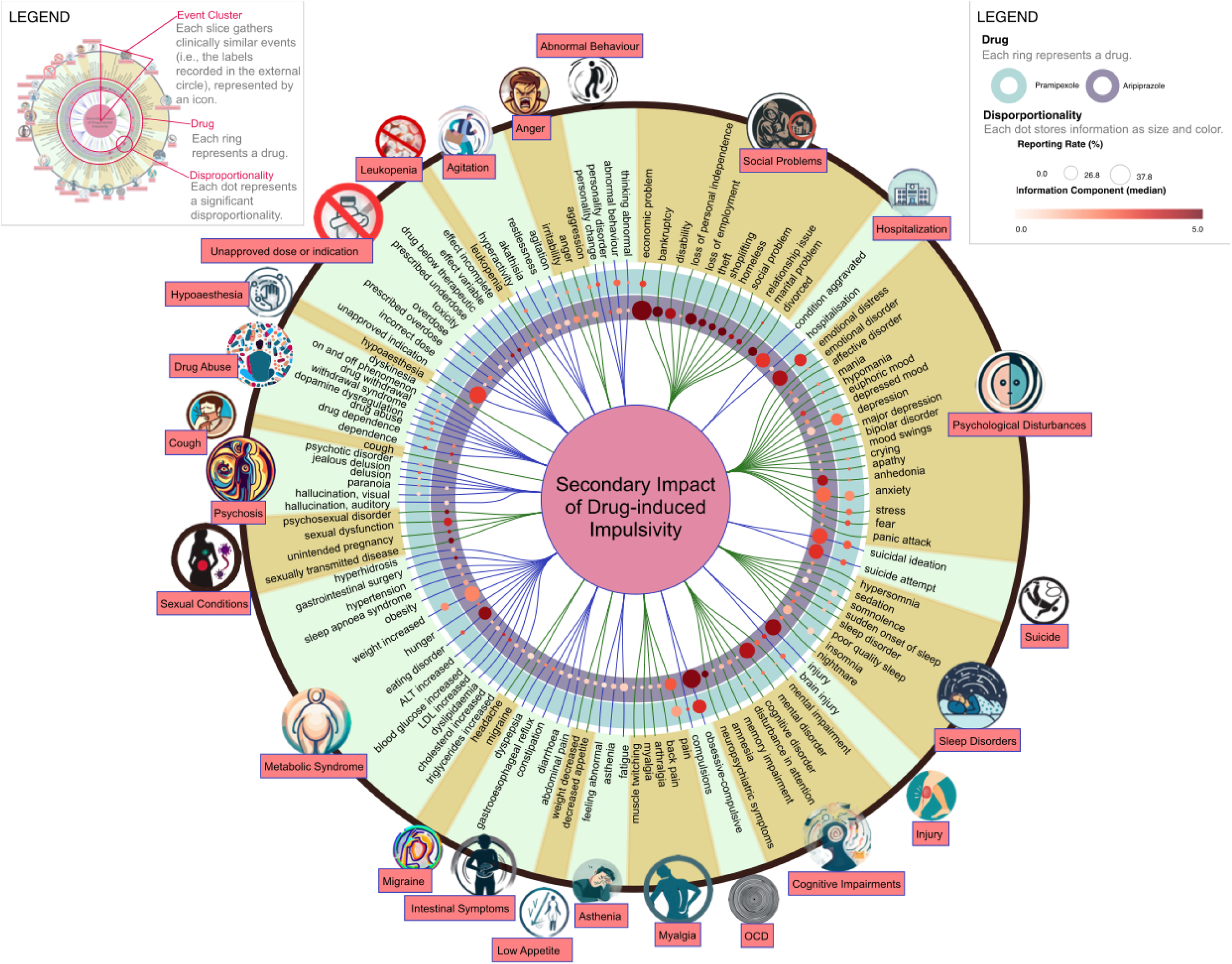
Secondary impact of drug-induced impulsivity. The dendrogram shows the events disproportionally reported with aripiprazole and pramipexole-related impulsivity. Events are gathered by clinical similarity in alternately colored slices, labeled on the outer border with a name and an icon. Disproportionalities are shown as dots organized in two colored rings, each representing a drug/case population. The dots’ size is proportional to the percent of reports showing the event, the color is darker for stronger disproportionality (higher median Information Component).

### 3.3 Network Analysis and sub-syndromes

In the second step we estimated the networks using three different approaches and identifying clusters of events. We included a total of 120 nodes (107 events disproportionally reported with impulsivity + 13 impulsivity PTs) and 70 nodes (56 + 14) for the aripiprazole and pramipexole network, respectively. Although the nodes remained constant, edges, clusters, and network properties were different in the three mathematical representations (**Table 3**, **S7, S10-S11, Figure S1-S8, S11-S16**). As expected, the degree centrality was highest for the most common events in Ising and for the rarest in PPMI. The Jaccard similarity was highest between the networks estimated by Ising and *ϕ* and lowest for the networks estimated by *ϕ* and PPMI (half of the links being different). The clustering was most overlapping between *ϕ*-PPMI, followed by *ϕ*-Ising and PPMI-Ising, as captured by the purity index. *ϕ* and PPMI tended to present clusters including multiple clusters from the Ising estimated network (Figures S11-S16).

**Table 3.**
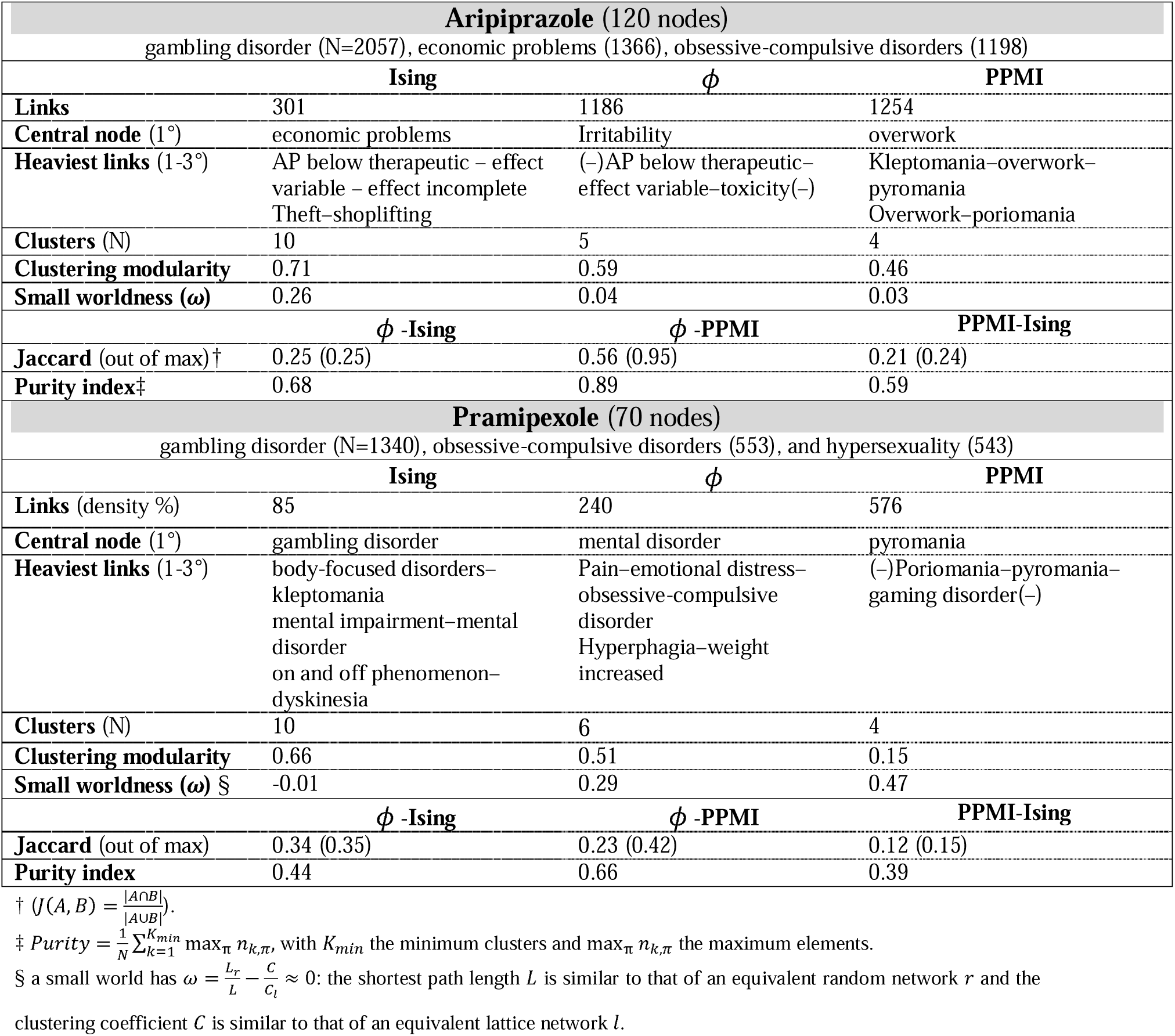
Network properties. The table shows the network properties for the three networks estimated for aripiprazole and pramipexole, respectively, and for their comparison. Legend: (–) means that there is also a strong link between the first and last element.

| **Aripiprazole (120 nodes)** |  |  |  |
| --- | --- | --- | --- |
| gambling disorder (N=2057), economic problems (1366), obsessive-compulsive disorders (1198) |  |  |  |
|  | **Ising** | **$\phi$** | **PPMI** |
| **Links** | 301 | 1186 | 1254 |
| **Central node (1°)** | economic problems | Irritability | overwork |
| **Heaviest links (1-3°)** | AP below therapeutic – effect variable – effect incomplete<br>Theft–shoplifting | (–)AP below therapeutic–effect variable–toxicity(–) | Kleptomania–overwork–pyromania<br>Overwork–poriomania |
| **Clusters (N)** | 10 | 5 | 4 |
| **Clustering modularity** | 0.71 | 0.59 | 0.46 |
| **Small worldness ($\omega$)** | 0.26 | 0.04 | 0.03 |
|  | **$\phi$ -Ising** | **$\phi$ -PPMI** | **PPMI-Ising** |
| **Jaccard (out of max)†** | 0.25 (0.25) | 0.56 (0.95) | 0.21 (0.24) |
| **Purity index‡** | 0.68 | 0.89 | 0.59 |
| **Pramipexole (70 nodes)** |  |  |  |
| gambling disorder (N=1340), obsessive-compulsive disorders (553), and hypersexuality (543) |  |  |  |
|  | **Ising** | **$\phi$** | **PPMI** |
| **Links (density %)** | 85 | 240 | 576 |
| **Central node (1°)** | gambling disorder | mental disorder | pyromania |
| **Heaviest links (1-3°)** | body-focused disorders–kleptomania<br>mental impairment–mental disorder<br>on and off phenomenon–dyskinesia | Pain–emotional distress–obsessive-compulsive disorder<br>Hyperphagia–weight increased | (–)Poriomania–pyromania–gaming disorder(–) |
| **Clusters (N)** | 10 | 6 | 4 |
| **Clustering modularity** | 0.66 | 0.51 | 0.15 |
| **Small worldness ($\omega$) §** | -0.01 | 0.29 | 0.47 |
|  | **$\phi$ -Ising** | **$\phi$ -PPMI** | **PPMI-Ising** |
| **Jaccard (out of max)** | 0.34 (0.35) | 0.23 (0.42) | 0.12 (0.15) |
| **Purity index** | 0.44 | 0.66 | 0.39 |

† $J(A, B) = \frac{|A \cap B|}{|A \cup B|}$ .
‡ $Purity = \frac{1}{N} \sum_{k=1}^{K_{min}} \max_{\pi} n_{k,\pi}$ , with $K_{min}$ the minimum clusters and $\max_{\pi} n_{k,\pi}$ the maximum elements.
§ a small world has $\omega = \frac{L_r}{L} - \frac{C}{C_l} \approx 0$ : the shortest path length $L$ is similar to that of an equivalent random network $r$ and the clustering coefficient $C$ is similar to that of an equivalent lattice network $l$ .

The three representations were considered as three different perspectives on the same phenomenon and composed into a single visualization for each population (**Figure 4, 5**). For a detailed description of the identified clusters, contextualized within the existing literature, see the discussion (paragraph 4.4).

**Figure 4.**
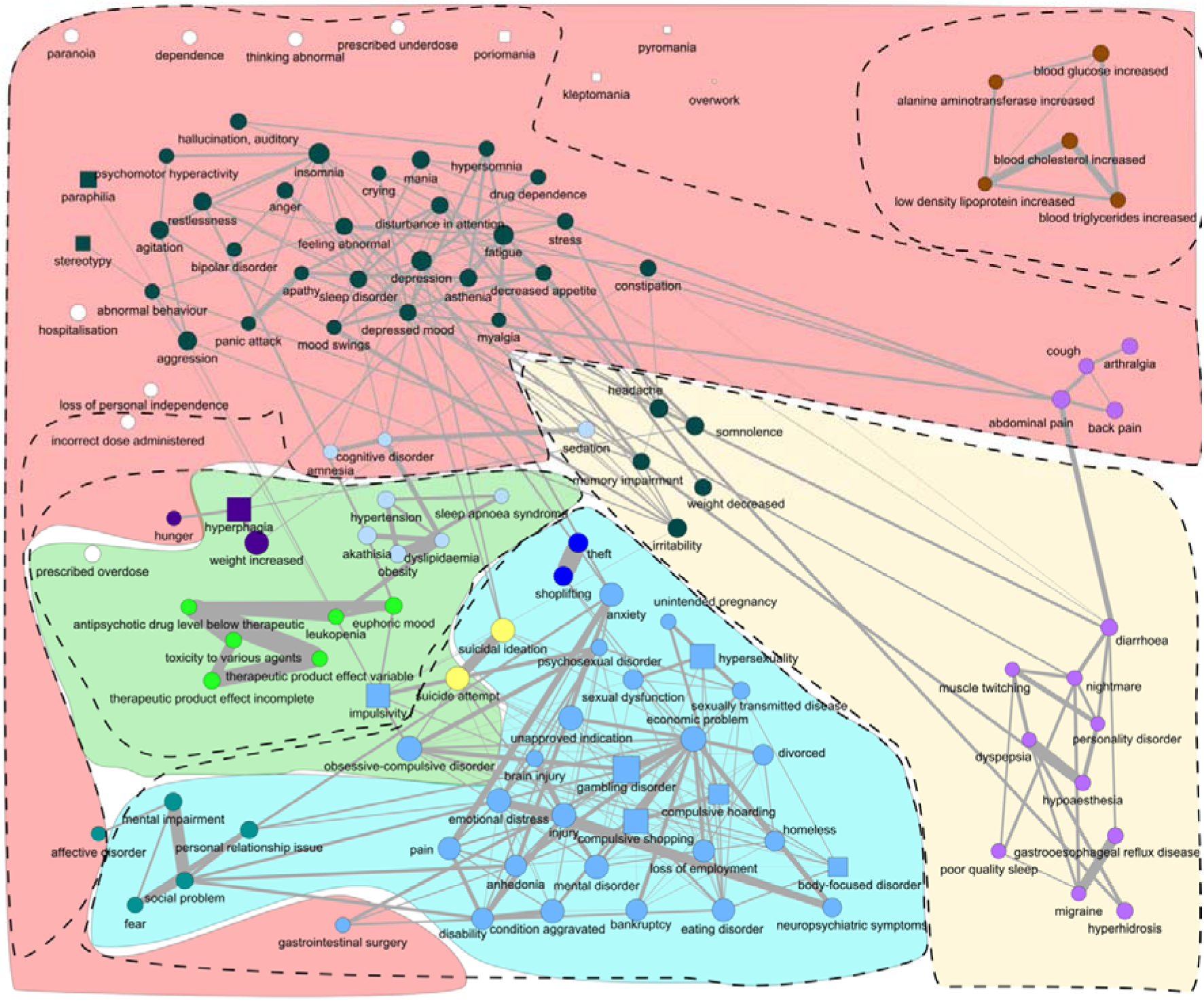
The secondary impact of aripiprazole-induced impulsivity. The network shows the events disproportionally reported with aripiprazole-related impulsivity and their pattern of co-reporting. Drug-induced impulsivity manifestations are shown as squares and other events as circles. Node colors identify clusters from the Ising estimation, dashed contours for the estimation, and colored contours for the PPMI estimation. The link width represents the weight of the links of the Ising, here chosen over the others because they are fewer and more conservative. The layout has been manually adjusted to reduce the overlapping. The layout calculated using a spring model with, as weight, the weights from the individual networks and the average of the weights of the three networks, after rescaling them from 0 to 1, is shown in the supplementary material.

**Figure 5.**
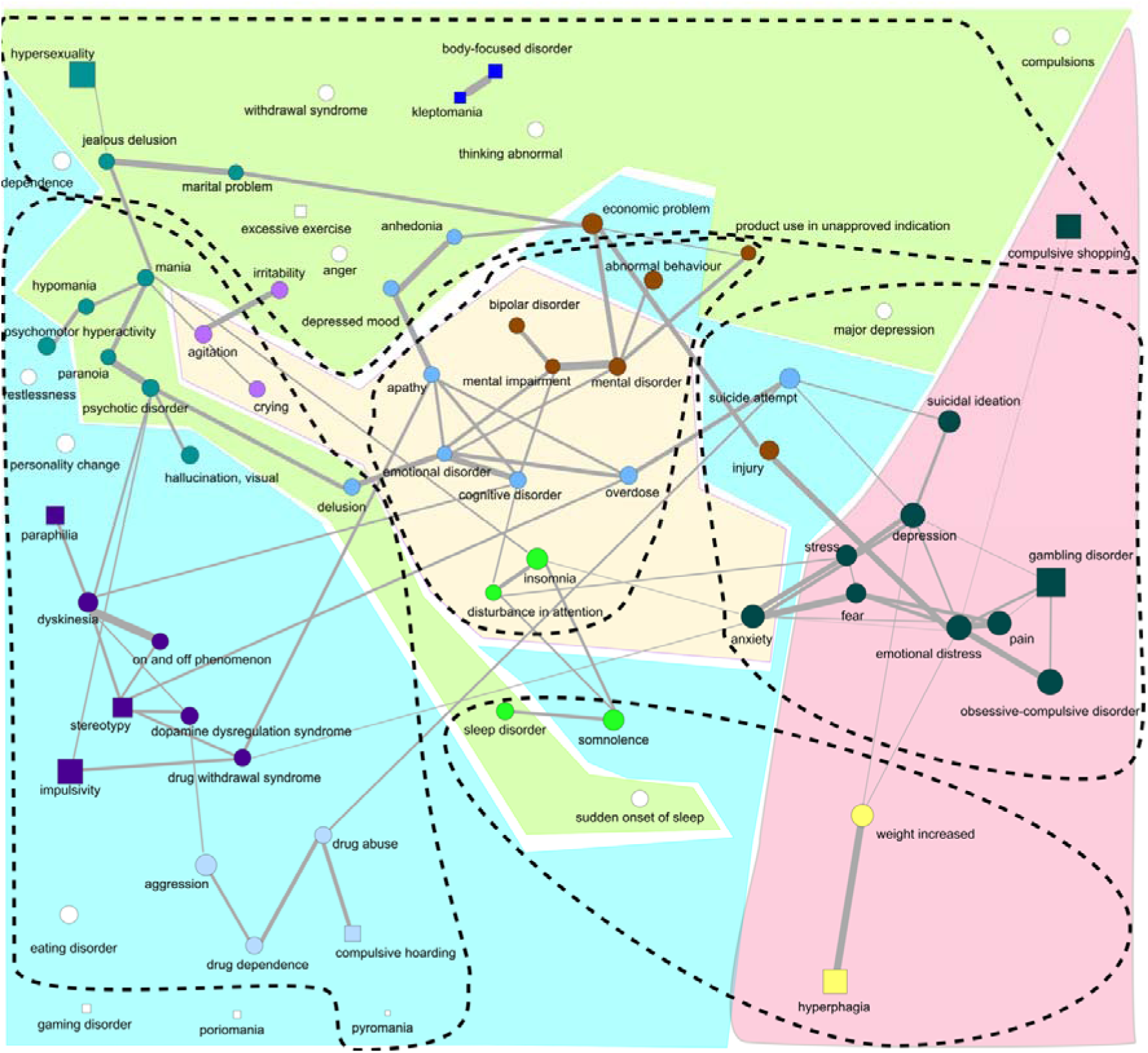
The secondary impact of pramipexole-induced impulsivity. The network shows the events disproportionally reported with pramipexole-related impulsivity and their pattern of co-reporting. Drug-induced impulsivity manifestations are shown as squares and other events as circles. Node colors identify clusters from the Ising estimation, dashed contours for the estimation, and colored contours for the PPMI estimation. The link width represents the weight of the links of the Ising, here chosen over the others because they are fewer and more conservative. The layout has been manually adjusted to reduce the overlapping. The layout calculated using a spring model with, as weight, the weights from the individual networks and the average of the weights of the three networks, after rescaling them from 0 to 1, is shown in the supplementary material.

3.4 Bayesian Network: The secondary impact of drug-induced impulsivity

The Bayesian Network yielded insights into the directional associations between co-reported events (see **Figure 6** and OSF repository [55]). High out-degree centrality identified pivotal events that likely heightened the likelihood of reporting other events (Figure S9-S10). Since this directed network only generates hypotheses, we preferred temporal terminology (i.e., preceding and following) to causal terminology even if no data on actual temporal sequences was included in the analyses.

**Figure 6.**
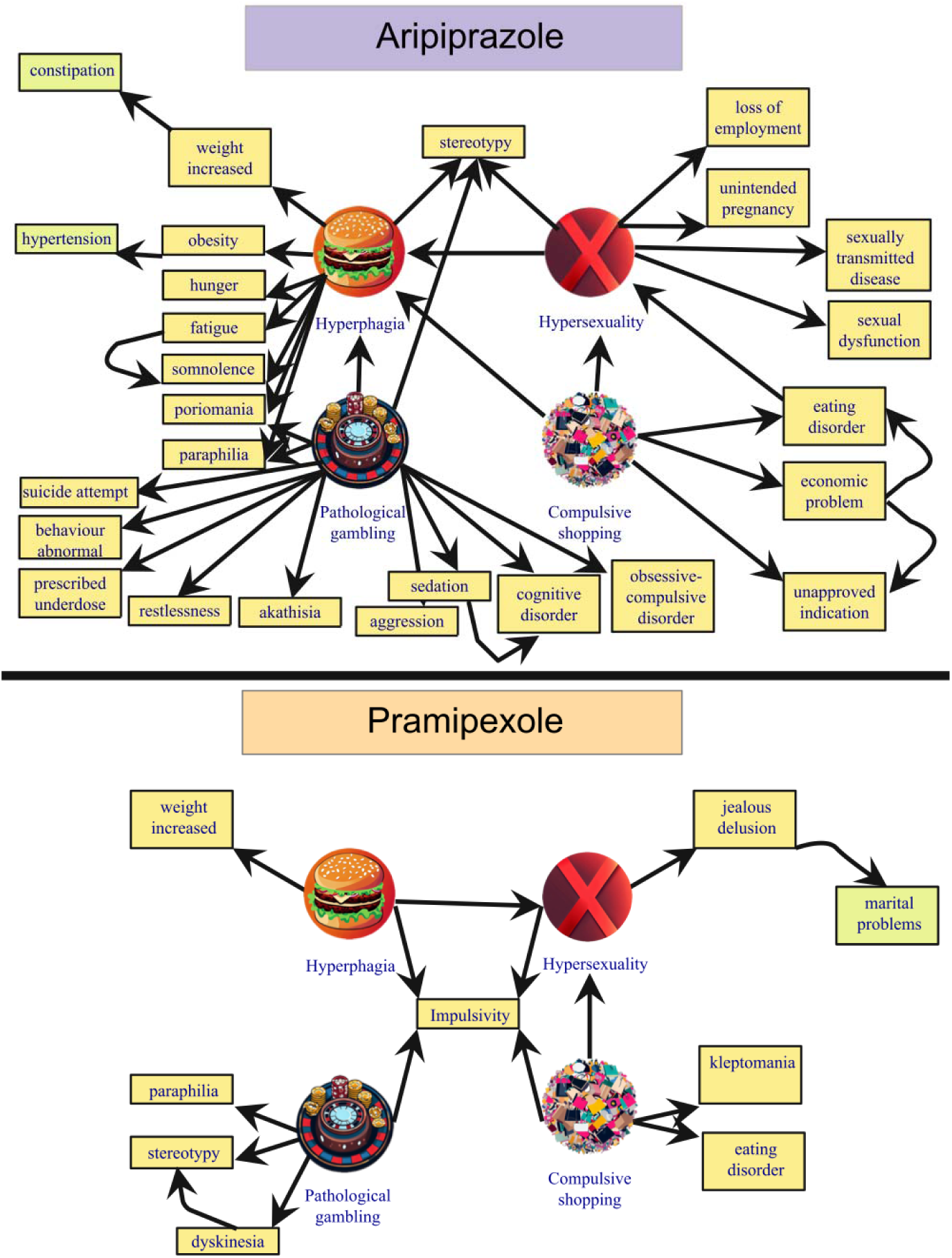
The secondary impact of the main drug-induced impulsivity, aripiprazole and pramipexole. The ego-networks extracted from the Bayesian Network show the potential direction of the co-reporting relationships between the events, thus providing insight into the direct and indirect impact of drug-induced impulsivity. Nodes linked to hyperphagia, hypersexuality, pathological gambling, and compulsive shopping are represented. Only out-neighbors of order equal or less than 1 are shown here, together with out-neighbors of order 2 considered relevant for clinical interpretation.

In pramipexole recipients, anxiety (3.55), emotional distress (2.92), and gambling (2.30) attained the highest out-degree centrality. Anxiety preceded insomnia (with irritability, somnolence, and attention disturbances), stress and depression (with suicide), fear, OCD, and emotional distress. Emotional distress preceded pain and injury (with major depression and economic problems), abnormal thinking and behavior, weight gain, and pathologic gambling. Furthermore, hypersexuality preceded delusional jealousy and marital difficulties, compulsive shopping stealing behaviors, and hyperphagia weight increase.

In aripiprazole recipients, economic problems (5.97), gambling (4.15), and hyperphagia (2.33) attained the highest out-degree centrality. Economic problems preceded theft, hoarding, divorce, loss of employment, homelessness, suicide, sex dysfunction, sexually transmitted diseases, and eating disorder. Gambling preceded aggressivity, suicide, cognitive disorders, hyperphagia, and paraphilia. Hyperphagia preceded somnolence and fatigue (with stress, attention disturbances, myalgia, cough), hunger, weight increase (with constipation), obesity (with hypertension), compulsive wandering, and paraphilic disorders. Anxiety preceded depression (with sleep disorders and suicide), fear and panic attacks (with relationship issues), pain and injury (with emotional distress, disability, anhedonia, and economic problems). Hypersexuality preceded sexual dysfunction, sexually transmitted diseases, unintended pregnancy, and loss of employment. Compulsive shopping preceded eating disorders and economic problems.

## 4. Discussion

### 4.1 Summary and Key Results

We investigated aripiprazole and pramipexole reports to capture the syndromes and sub-syndromes related to drug-induced impulsivity.

The event-event disproportionality analysis revealed signs and symptoms commonly reported alongside impulsivity, thus delineating an impulsivity syndrome, separately for aripiprazole and pramipexole recipients. The impulsivity syndrome encompassed mainly psychosocial events but also organic conditions.

The network analysis identified meaningful clusters, such as delusional jealousy (also known as Othello syndrome [56]) and dopamine dysregulation syndrome (i.e., the excessive use of levodopa) in pramipexole recipients, and obesity-hypoventilation syndrome (historically Pickwickian syndrome) and social issues in aripiprazole recipients. In particular, employing more sensitive network analyses like Phi and PPMI generated more interconnected networks, which identified potential macro-clusters combining several smaller clusters identified by the Ising model.

Crucially, potential causal mechanisms and secondary consequences of drug-induced impulsivity can be highlighted by Bayesian Network methods, providing targets for potential interventions. Anxiety and economic problems emerged as pivotal events that could be potentially targeted to disrupt the chain of events and alleviate the burden of drug-induced impulsivity: for instance, monitoring and effectively managing anxiety or providing financial guidance or legal guardianship to prevent wasteful spending. Since marital problems affect caregivers’ QoL and increase the risk of early placement in nursing homes [22], addressing delusional jealousy and economic problems, identified as factors preceding marital problems, may be critical for preserving wellbeing in pramipexole recipients.

In order to better contextualize and qualify these findings, we now discuss in detail the results of case retrieval, disproportionality analysis, network analysis, Bayesian network, and the limits and conclusions of the study.

### 4.2 Case Retrieval

Our findings align with established risk factors, including male gender and younger age [57,58], Parkinson’s Disease (PD) [59,60] and depression [61]. Commonly reported impulsivity manifestations included the “four knights”[9] (i.e., gambling, shopping, hyperphagia, and hypersexuality), garnering special attention due to their pronounced impact on QoL. Other manifestations were body-focused repetitive behaviors, paraphilic disorders, and hoarding.

### 4.3 Disproportionality analysis: the drug-induced impulsivity syndrome

By performing the event-event disproportionality analysis within each drug population separately, comparing reports involving impulsivity with those encompassing various reactions – other than impulsivity – to the same drug, we addressed indication bias and other confounding factors. This comparative analysis served as a rigorous filter, allowing us to sift through the complex data and unveil the genuine characteristics associated with impulsivity, as well as those arising from the dynamic interaction between impulsivity and the underlying drug or disease, excluding traits tied solely to the underlying drug or disease.

This approach revealed a complex syndrome, characterized by psychosocial, cognitive, psychosomatic, and metabolic events. The syndromes identified for impulsivity within pramipexole and aripiprazole recipients differ significantly. Multiple factors may contribute to the seemingly higher burden on QoL observed with aripiprazole-induced impulsivity, with functional (or psychosomatic) manifestations and social issues impacting work, relationships, and economics. Pramipexole is primarily administered to older patients with hypodopaminergic conditions, characterized by motor impairment and reduced motivational drive. These patients, well managed and supported by caregivers because of the later onset and clear neurologic origin of the disease, may experience a mitigated drug-induced impulsivity burden. Conversely, aripiprazole is prescribed to younger patients with mood and psychotic disorders, often linked to hyperdopaminergic states and a pre-existing diathesis for impulsivity. Challenges for caregivers and social support are heightened in these cases due to earlier onset, psychiatric origins, and stigma, potentially leading to a greater burden. Further, over a third of aripiprazole cases were submitted by lawyers. This may be explained either by a potentially malicious overreporting for legal compensation (cfr., Abilify lawsuit) [62] or by a reaction to underdiagnosis by physicians, who may be hesitant to attribute behavioral changes to the medication when underlying psychiatric conditions are present. In reports filed by lawyers, the desire to win legal compensation may have prompted more detailed descriptions of the impact of impulsivity on QoL, or possibly even exaggerated the impact, thus contributing to the observed differences between the two drugs. Intriguingly, there could also be an ascertainment bias, as neurologists prescribing pramipexole may be less aware of psychiatric issues compared to psychiatrists prescribing aripiprazole.

### 4.4 Network Analysis: sub-syndromes

Network analysis in pharmacovigilance, complementary to other unsupervised approaches such as vigiGroup [63], is a promising tool to detect potential syndromes and subsyndromes, to help the characterization of signals. Employing three estimation methods, the network analysis revealed potential subsyndromes associated with specific impulsivity expressions in the two populations (**Figure 7**). The Ising delineated well-defined clusters, while PPMI and *ϕ* emphasized inter-clusters relationships.

**Figure 7.**
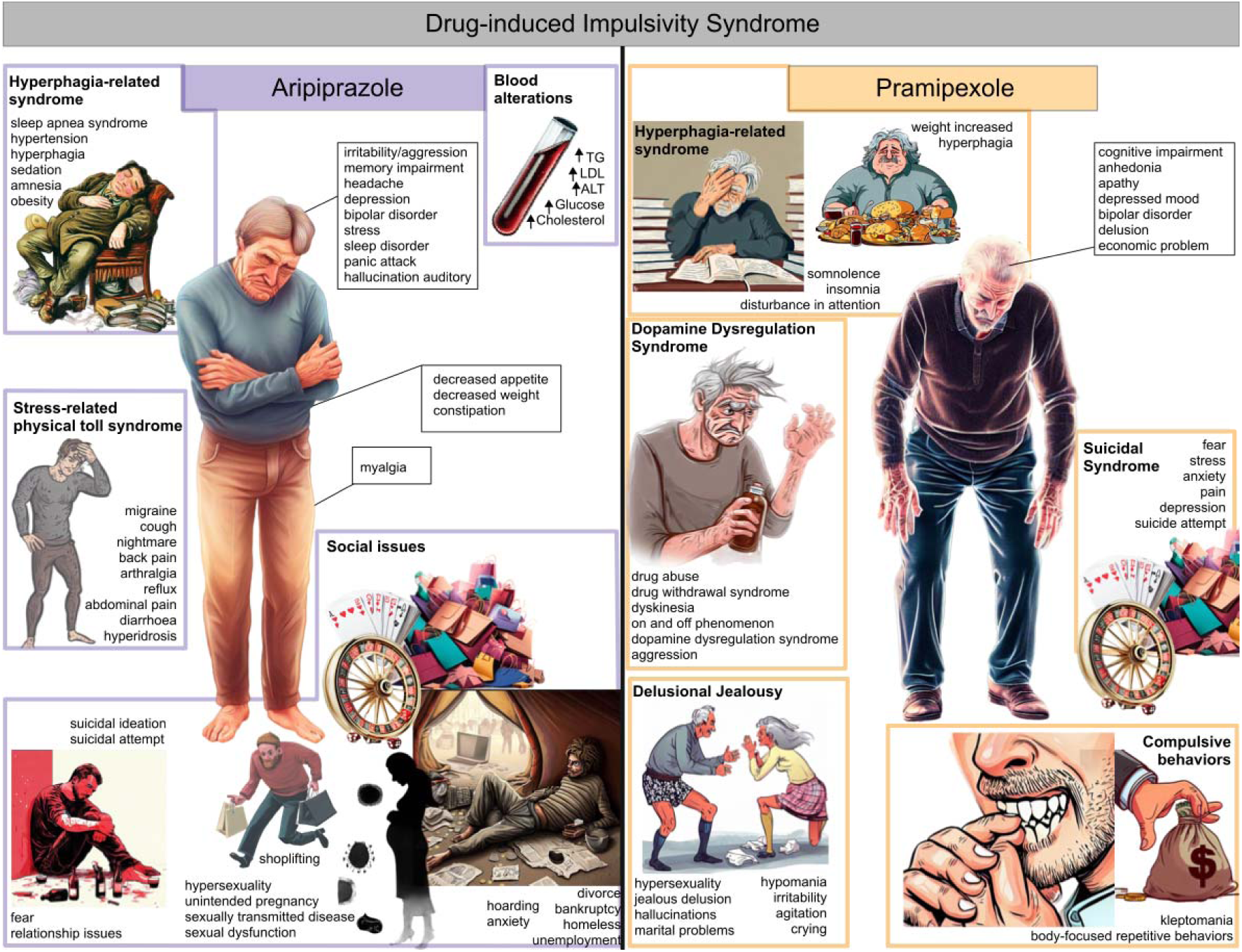
Drug-induced impulsivity syndrome, aripiprazole and pramipexole. The main syndrome, representing one or more strongly interconnected central clusters of symptoms and signs identified through network analysis, is depicted as the central figure. Other potential sub-syndromes are shown on the sides highlighted with a colored square.

By incorporating various expressions of impulsivity, we anticipated that the central cluster would encompass the key features of impulsivity regardless of its form, whether they act as risk factors or consequences of impulsivity. In both populations, cognitive and mood disorders (e.g., cognitive and memory impairment, bipolar disorder, depression) were included in the central cluster. Notably, they have been recorded as frequently associated with drug-induced impulsivity and contributing to disability development [64]. Obesity-hypoventilation syndrome [65], involving weight gain, cognitive and sleep disorders, and sedation, was consistent in both populations but seemingly heavier in aripiprazole recipients, which also reported obesity, sleep apnoea syndrome, hypertension, and metabolic blood alterations (increased lipids, transaminases, and glucose in the blood), supporting the observed link between hyperphagia and diabetes onset [66].

For aripiprazole recipients (Figure 4), the central cluster also included panic attack and auditory hallucinations, sleep disorders, decreased appetite, and stress. Stress was further connected to a psychosomatic sub-syndrome involving irritability, migraine, back and abdominal pain, reflux, diarrhoea, constipation, and hyperidrosis. Gambling and shopping were linked to pervasive social issues (hoarding, unemployment, homeless, bankruptcy, divorce), theft, and suicidal ideation and attempts (already observed during hyperdopaminergic impulsive states [67]). Hypersexuality was linked to unintended pregnancy, sexually transmitted diseases, and sexual dysfunction.

Among pramipexole recipients (Figure 5), the central cluster also included apathy, delusion, and economic problems. The dopamine dysregulation sub-syndrome (a manifestation of pathological impulsivity marked by excessive levodopa use [68–70], co-administered with dopamine agonists to better control motor symptoms), involved on and off phenomenon (oscillations in effectiveness and motor and motivational symptoms), excessive levodopa use to avoid off phases, and dopamine agonist withdrawal syndromes (DAWS) upon discontinuation [71]. A cluster aligned with paranoid delusional jealousy (false and unwavering belief in the partner’s unfaithfulness), often seen in PD with drug-induced hypersexuality [72] and here characterized by delusional jealousy, hallucinations, irritability, crying, and marital problems. Notably, marital problems have been associated with early placement in nursing homes and worse prognosis [73]. We also found a cluster with fear, pain, stress, anxiety, depression, and suicidal ideation, indicative of the transformation of reward-driven impulsivity into stressful risk-averting compulsivity over time [74]. Finally, the co-reporting of two archetypal compulsive symptoms—body-focused repetitive behaviors and stealing behaviors—was evident.

### 4.5 Bayesian Network: the secondary impact of drug-induced impulsivity

This interplay of events within the context of drug-induced impulsivity is intricate and multifaceted. Events reported alongside drug-induced impulsivity may result from impulsivity itself (like financial problems from gambling) or predispose individuals to impulsivity (e.g., bipolar disorder). Sometimes, events can both trigger and be exacerbated by drug-induced impulsivity (e.g., anxiety [75,76]). Sometimes events are concomitantly mentioned for precision, such as in cases of semantic overlap (e.g., theft and shoplifting, or injury and brain injury). Events associated with drug-induced impulsivity may even be synonyms for well-known impulsivity expressions (e.g., restlessness, referring to excessive wandering and poriomani), or could be the very reason for prescribing the drug, as seen in the off-label use of aripiprazole to prevent behavioral and cognitive decline in brain injury [77] or to address drug dependence [78,79]. In order to get insights into potential directional associations to attempt the formulation about clinically plausible causal sequences, we implemented a Bayesian Network. Anxiety emerged as a central factor, preceding insomnia, irritability, cognitive impairment, stress, injury, pain (linked to disability and economic problems), depression, and even suicidal ideation. Drug-induced impulsivity manifestations appeared to exacerbate each other. Economic problems had the highest out-degree centrality among aripiprazole recipients, preceding theft, relationship difficulties, and suicidal ideation. The Bayesian Network provides clinicians with valuable insights on the pivotal nodes that could be targeted by interventions to disrupt the cascade of events and ameliorate the secondary impact of drug-induced impulsivity. It also highlighted secondary ramifications of main impulsivity manifestations: hypersexuality precedes marital problems through delusional jealousy in pramipexole recipients, while it precedes unintended pregnancy and sexually transmitted diseases in aripiprazole recipients; hyperphagia precedes weight increase in pramipexole recipients and obesity, somnolence, and cognitive impairment in aripiprazole. Marital problems, following delusional jealousy and economic problems in pramipexole recipients, may be of particular interest since they are associated with an early placement in nursing homes [73]. Finally, the Bayesian Network seems to support the higher secondary impact of drug-induced impulsivity in aripiprazole recipients.

### 4.6 Limitations and Further Developments

While this study provides valuable insights into the intricate interplay of events related to drug-induced impulsivity and its subsequent implications, it is crucial to acknowledge its limitations.

Spontaneous reports, while uniquely granting access to patients’ perspective, are susceptible to biases like under-reporting, missing data, and unverified reliability, preventing reliable incidence or prevalence estimates. The high contribution of reports from lawyers may have influenced the higher psychosocial impact attributed to aripiprazole-induced impulsivity. Nonetheless, this study sets the foundation for further studies and a potential score to assess the impact of ADRs on QoL.

Limitations in network analysis methodologies adopted include the Ising estimation’s assumptions (pairwise interaction, linear effects, and binary variables), and the inability to account for time and severity in symptom manifestation. The incorporation of negative links could facilitate a more nuanced separation of symptoms that infrequently co-occur. The Bayesian Network lacks bidirectional relationships and cyclic feedback loops and would require the inclusion of all shared causes between any two events (causal Markov condition) to achieve its full capacity to illuminate causality. These limitations could be rectified by integrating clinical longitudinal data and embedding temporal aspects into the network analysis.

Looking ahead, a broader definition of drug-induced impulsivity could improve sensitivity in case retrieval. Conditions like suicide attempts, hypersomnia, obsessive-compulsive symptoms, explosive anger, personality changes, disturbance in attention, and drug dependence might represent different expressions of this underdefined condition, warranting further exploration [10,70].

### 4.7 Conclusion

The profound impact of drug-induced impulsivity reverberates across patients and their families, encompassing psychosocial challenges and organic complications such as metabolic syndrome (in the case of hyperphagia), and sexual health issues (in the case of hypersexuality). Recognizing these potential consequences is crucial for informed pharmacological management and diligent patient monitoring. Network analysis has revealed intriguing co-reporting patterns among adverse events, leading to their classification as sub-syndromes. Notable examples include the emergency of obesity-hypoventilation syndrome with hyperphagia and associations of hypersexuality with delusional jealousy in pramipexole recipients and unintended pregnancy and sexually transmitted diseases in aripiprazole recipients. Our parallel approach effectively avoids the risk of disease-related diathesis compromising analytical integrity, enhancing the robustness of our findings.

For clinicians, this study emphasizes the potential burden of drug-induced impulsivity, and therefore the necessity of meticulous scrutiny into patients’ medical histories. Factors such as age, gender, pre-existing mood disorders and family history should be red flags, warranting heightened vigilance. While transitioning to an alternative active ingredient may mitigate impulsivity, it is not always feasible or adequate. Monitoring for potential complications, as unveiled in our work (e.g., obesity-hypoventilation syndrome and delusional jealousy), is pivotal when such transitions are not a viable solution. For example, an overlooked delusional jealousy may result in marital problems and early nursing home placement.

Central to our findings is the pivotal realization that drug reactions rarely occur in isolation; instead, they manifest as syndromes with diverse signs and symptoms. These can be direct reactions to the drug itself, secondary consequences to the reaction, risk factors for the reaction, or comorbidities. Causal chains and loops can contribute to symptom aggravation and chronicity. Identifying syndromes and sub-syndromes, combining network strategies with traditional techniques and clinical judgment, proves a potent strategy for delving into the secondary impact of adverse drug reactions and fostering heightened awareness within clinical practice.

In sum, the intricate relationships between signs and symptoms, coupled with the insights from the Bayesian Network, underscore the multifaceted nature of drug-induced impulsivity. More significantly, it equips research, and secondarily clinicians, with indispensable tools to identify potential intervention points, decipher causal sequences, and mitigate the cascading secondary effects associated with drug-induced impulsivity. In doing so, this study contributes to advancing our comprehension and management of drug-induced impulsivity, ultimately enhancing the well-being and care of affected patients.

## Data Availability

The data we used comes from the FDA Adverse Event Reporting System (FAERS), and is made publicly available by the FDA as quarterly data downloadable at https://fis.fda.gov/extensions/FPD-QDE-FAERS/FPD-QDE-FAERS.html. The algorithm for cleaning FAERS data is open-source at https://github.com/fusarolimichele/DiAna, and the cleaned database is available on an OSF repository and through the R package DiAna.

https://osf.io/k9v6s/

## Acknowledgements

MedDRA®, version 25.0 was developed under the auspices of the International Council for Harmonization of Technical Requirements for Pharmaceuticals for Human Use.

Part of the work was presented orally at the SIF congress, 2023, in Rome.

Icons for figures 1, 2, 6, and 7 were obtained using Bing Image Creator.

## Declarations

## Ethics approval

not applicable because spontaneous reports of the FAERS are anonymous and publicly available.

## Consent to participate

not applicable because spontaneous reports of the FAERS are anonymous and publicly available.

## Consent for publication

not applicable because spontaneous reports of the FAERS are anonymous and publicly available.

## Software used

Analyses were performed using R (version 4.2.1)[80] and Python (version 3.8.16)[81]. We relied on several essential packages for network analysis: *IsingFit*[47], *igraph*[82], *psych*[83], and *bnlearn*[35].

## Availability of data and material

The data we used comes from the FDA Adverse Event Reporting System (FAERS), and is made publicly available by the FDA as quarterly data downloadable at https://fis.fda.gov/extensions/FPD-QDE-FAERS/FPD-QDE-FAERS.html. The algorithm for cleaning FAERS data is open-source at https://github.com/fusarolimichele/DiAna, and the cleaned database is available on an OSF repository[84] and through the R package DiAna[85].

## Code availability

The code for the project is available on an OSF repository[55], the function for the Ising network is also available in the DiAna package (cfr. “network_analysis()”)[85].

## Author contributions

MF, SP, LM, conceptualized and designed the study. MF, SP, LM developed the methodology. The formal analysis was performed by MF, SP, LM, VG. MF, SP, VG performed the visualization. MF, SP, LM, VG wrote the original draft. All the authors strongly contributed to the interpretation of results, and to the review and editing of the draft. All the authors read and approved the final version.

