## Supplementary Material for "Unveiling the Burden of Drug-Induced Impulsivity: A Network Analysis of the FDA Adverse Event Reporting System"

**Table S1. Expressions of pathological impulsivity.** MedDRA query and behavioral subqueries used for retrieving drug-induced impulsivity.

| Impulsivity expression | Preferred Terms (PT) |
|---|---|
| Pathological gambling | gambling disorder, gambling |
| Hypersexuality | compulsive sexual behaviour, hypersexuality, excessive masturbation, excessive sexual fantasies, libido increased, sexual activity increased, kluver-bucy syndrome |
| Paraphilia | erotophonophilia, exhibitionism, fetishism, frotteurism, masochism, paraphilia, pedophilia, sadism, transvestism, voyeurism, sexually inappropriate behaviour |
| Compulsive shopping | compulsive shopping |
| Hyperphagia | binge eating, food craving, hyperphagia, increased appetite |
| Gaming disorder | gaming disorder |
| Pyromania | pyromania |
| Kleptomania | kleptomania |
| Compulsive hoarding | compulsive hoarding |
| Excessive exercise | excessive exercise |
| Overwork | overwork |
| Poriomania | poriomania |
| Body-focused repetitive behaviors | compulsive cheek biting, compulsive lip biting, dermatillomania, dermatophagia, nail picking, compulsive handwashing, trichotemnomania, trichotillomania, onycophagia, thumb sucking |
| Stereotypy | automatism, stereotypy |
| Impulsivity | impulse-control disorder, impulsive behaviour, disinhibition, behavioural addiction |

**Table S2. Manifestation of impulsivity in the two populations investigated.**

| Pramipexole | | Aripiprazole | |
|---|---|---|---|
| Gambling disorder | 1345 (4.87%) | Gambling disorder | 2067 (2.58%) |
| Hypersexuality | 612 (2.22%) | Hypersexuality | 1077 (1.34%) |
| Impulsivity | 453 (1.64%) | Compulsive shopping | 1029 (1.28%) |
| Compulsive shopping | 384 (1.39%) | Hyperphagia | 868 (1.08%) |
| Hyperphagia | 334 (1.21%) | Impulsivity | 730 (0.91%) |
| Stereotypy | 87 (0.32%) | Body-focused disorder | 316 (0.39%) |
| Paraphilia | 64 (0.23%) | Compulsive hoarding | 244 (0.30%) |
| Compulsive hoarding | 26 (0.09%) | Paraphilia | 78 (0.10%) |
| Body-focused disorder | 18 (0.07%) | Stereotypy | 42 (0.05%) |
| Excessive exercise | 7 (0.03%) | Poriomania | 15 (0.02%) |
| Kleptomania | 7 (0.03%) | Kleptomania | 6 (0.01%) |
| Gaming disorder | 3 (0.01%) | Pyromania | 4 (0.00%) |
| Poriomania | 2 (0.01%) | Overwork | 1 (0.00% |
| Pyromania | 1 (0.00%) | | |

**Table S3. Features of pramipexole reports.** We compared cases of pramipexole-related pathological impulsivity vs any other report recording pramipexole. Chi-square test was performed for categorical values, Mann-Whitney for continuous ones, and p-values were corrected for multiple comparison using the Holm-Bonferroni method.

| VARIABLE | Impulsivity Reports N = 2,066 | | Other Reports N = 25,535 | | p-value |
|---|---|---|---|---|---|
| | N | % | N | % | |
| **Gender** | | | | | **<0.001** |
| Woman | 821 | 42.58 | 14,964 | 63.01 | |
| Man | 1,107 | 57.42 | 8,784 | 36.99 | |
| Unknown | 138 | - | 1,787 | - | |
| **Age (years old)** | | 56 (48-64) | | 67 (57-75) | **<0.001** |
| Unknown | | 903 | | 8,482 | |
| **Weight (Kg)** | | 83 (72-95) | | 79 (65-95) | **<0.001** |
| Unknown | | 1,386 | | 16,330 | |
| **Outcome** | | | | | **<0.001** |
| Death | 40 | 1.94 | 1,883 | 7.37 | |
| Life Threatening | 50 | 2.42 | 713 | 2.79 | |
| Disability | 90 | 4.36 | 687 | 2.69 | |
| Required Intervention | 27 | 1.31 | 74 | 0.29 | |
| Hospitalization | 305 | 14.76 | 7,378 | 28.89 | |
| Congenital | 0 | 0.00 | 20 | 0.08 | |
| Other Serious | 627 | 30.35 | 6,205 | 24.30 | |
| Non-Serious | 927 | 44.87 | 8,575 | 33.58 | |
| **Reporter** | | | | | **<0.001** |
| Consumer | 1,092 | 56.73 | 10,315 | 44.75 | |
| Healthcare practitioner | 50 | 2.60 | 1,378 | 5.98 | |
| Lawyer | 29 | 1.51 | 220 | 0.95 | |
| Other | 380 | 19.74 | 3,366 | 14.60 | |
| Pharmacist | 29 | 1.51 | 1,822 | 7.90 | |
| Physician | 345 | 17.92 | 5,950 | 25.81 | |
| Unknown | 141 | - | 2,484 | - | |

**Table S4. Features of aripiprazole reports.** We compared cases of aripiprazole-related pathological impulsivity vs any other report recording aripiprazole. Chi-square test was performed for categorical values, Mann-Whitney for continuous ones, and p-values were corrected for multiple comparison using the Holm-Bonferroni method.

| VARIABLE | | Impulsivity Reports N = 3,609 | | Other Reports N = 76,629 | | **p-value** |
|---|---|---|---|---|---|---|
| | | N | % | N | % | |
| **Gender** | | | | | | **<0.001** |
| | Woman | 1,624 | 51.41 | 40,454 | 59.28 | |
| | Man | 1,535 | 48.59 | 27,792 | 40.72 | |
| | Unknown | 450 | - | 8,383 | - | |
| **Age (years old)** | | | 40 (25-51) | | 41 (26-55) | **<0.001** |
| | Unknown | | 1,951 | | 25,267 | |
| **Weight (Kg)** | | | 82 (68-104) | | 79 (63-98) | **<0.001** |
| | Unknown | | 2,876 | | 55,594 | |
| **Outcome** | | | | | | **<0.001** |
| | Death | 31 | 0.86 | 4,099 | 5.35 | |
| | Life Threatening | 61 | 1.69 | 2,823 | 3.68 | |
| | Disability | 483 | 13.38 | 1,868 | 2.44 | |
| | Required Intervention | 12 | 0.33 | 280 | 0.37 | |
| | Hospitalization | 1,205 | 33.39 | 17,921 | 23.39 | |
| | Congenital | 0 | 0.00 | 325 | 0.42 | |
| | Other Serious | 896 | 24.83 | 21,051 | 27.47 | |
| | Non-Serious | 921 | 25.52 | 28,262 | 36.88 | |
| **Reporter** | | | | | | **<0.001** |
| | Consumer | 1,416 | 40.18 | 34,977 | 48.55 | |
| | Healthcare practitioner | 173 | 4.91 | 4,146 | 5.76 | |
| | Lawyer | 1,201 | 34.08 | 790 | 1.10 | |
| | Other | 256 | 7.26 | 10,795 | 14.98 | |
| | Pharmacist | 45 | 1.28 | 4,159 | 5.77 | |
| | Physician | 433 | 12.29 | 17,173 | 23.84 | |
| | Unknown | 85 | - | 4,589 | - | |

**Table S5. Events disproportionally reported with aripiprazole-induced impulsivity**. Ordered by N.

| Event | Perc (%) | Information Component Median (95%CI) [N] |
|---|---|---|
| economic problem | 37,85 | 4.28 (4.19-4.34) [1366] |
| obsessive-compulsive disorder | 33,19 | 4.16 (4.07-4.23) [1198] |
| product use in unapproved indication | 27,51 | 2.8 (2.69-2.87) [993] |
| Injury | 25,10 | 4 (3.9-4.08) [906] |
| mental disorder | 23,75 | 3.7 (3.59-3.78) [857] |
| suicidal ideation | 23,47 | 2.69 (2.58-2.77) [847] |
| weight increased | 23,19 | 1.83 (1.71-1.91) [837] |
| emotional distress | 22,80 | 3.75 (3.63-3.83) [823] |
| Anxiety | 21,56 | 2.2 (2.09-2.29) [778] |
| suicide attempt | 21,50 | 2.99 (2.88-3.08) [776] |
| condition aggravated | 20,34 | 2.91 (2.79-3) [734] |
| eating disorder | 16,15 | 4.05 (3.92-4.15) [583] |
| Pain | 14,35 | 2.45 (2.3-2.55) [518] |
| loss of employment | 12,64 | 4.33 (4.18-4.44) [456] |
| Disability | 11,42 | 3.79 (3.63-3.91) [412] |
| Anhedonia | 10,94 | 3.75 (3.59-3.87) [395] |
| Bankruptcy | 10,58 | 4.43 (4.26-4.55) [382] |
| Insomnia | 8,04 | 1.13 (0.94-1.27) [290] |
| Divorced | 7,59 | 4.38 (4.19-4.53) [274] |
| Homeless | 6,93 | 4.37 (4.16-4.52) [250] |
| sexual dysfunction | 6,79 | 3.25 (3.04-3.4) [245] |
| Depression | 6,62 | 0.78 (0.57-0.94) [239] |
| Fatigue | 6,62 | 0.78 (0.57-0.94) [239] |
| Theft | 5,79 | 4.32 (4.09-4.48) [209] |
| Shoplifting | 5,02 | 4.37 (4.12-4.54) [181] |
| neuropsychiatric symptoms | 4,74 | 4.35 (4.1-4.53) [171] |
| Irritability | 4,54 | 1.73 (1.47-1.91) [164] |
| Aggression | 4,35 | 1.26 (1-1.45) [157] |
| Akathisia | 4,05 | 0.89 (0.62-1.09) [146] |
| Somnolence | 4,02 | 0.38 (0.1-0.57) [145] |
| Headache | 4,02 | 0.51 (0.24-0.71) [145] |
| Restlessness | 3,69 | 1.08 (0.79-1.28) [133] |
| Agitation | 3,57 | 0.76 (0.47-0.97) [129] |
| abnormal behaviour | 3,44 | 1.28 (0.98-1.49) [124] |
| Mania | 3,13 | 1.11 (0.8-1.33) [113] |
| mental impairment | 3,08 | 2.26 (1.95-2.49) [111] |
| sexually transmitted disease | 3,05 | 4.24 (3.93-4.47) [110] |
| Hypertension | 2,96 | 1.01 (0.69-1.24) [107] |
| Asthenia | 2,96 | 0.89 (0.57-1.13) [107] |
| personal relationship issue | 2,94 | 3.9 (3.58-4.13) [106] |
| Obesity | 2,91 | 1.42 (1.1-1.66) [105] |
| weight decreased | 2,72 | 0.79 (0.46-1.03) [98] |
| disturbance in attention | 2,72 | 1.51 (1.17-1.75) [98] |
| feeling abnormal | 2,72 | 0.37 (0.04-0.61) [98] |
| euphoric mood | 2,69 | 3.02 (2.68-3.26) [97] |
| social problem | 2,66 | 3.78 (3.45-4.03) [96] |
| memory impairment | 2,60 | 1.05 (0.71-1.29) [94] |
| sleep disorder | 2,60 | 1.53 (1.19-1.77) [94] |
| Diarrhoea | 2,49 | 0.54 (0.19-0.8) [90] |
| therapeutic product effect incomplete | 2,38 | 1.88 (1.52-2.14) [86] |
| depressed mood | 2,36 | 1.68 (1.32-1.94) [85] |
| brain injury | 2,36 | 3.11 (2.75-3.37) [85] |
| Hyperhidrosis | 2,27 | 0.91 (0.54-1.17) [82] |

| Event | Perc (%) | Information Component Median (95%CI) [N] |
|---|---|---|
| sedation | 2,24 | 0.72 (0.36-0.99) [81] |
| anger | 2,19 | 1.39 (1.02-1.66) [79] |
| hallucination, auditory | 2,13 | 0.63 (0.25-0.9) [77] |
| prescribed overdose | 2,13 | 1.89 (1.52-2.17) [77] |
| constipation | 2,11 | 0.83 (0.45-1.1) [76] |
| back pain | 2,11 | 1.12 (0.74-1.39) [76] |
| blood glucose increased | 2,08 | 0.73 (0.34-1) [75] |
| psychosexual disorder | 2,02 | 4.1 (3.71-4.38) [73] |
| stress | 2,00 | 1.9 (1.51-2.18) [72] |
| hospitalisation | 2,00 | 0.68 (0.29-0.96) [72] |
| blood cholesterol increased | 1,94 | 0.95 (0.55-1.23) [70] |
| arthralgia | 1,88 | 0.78 (0.37-1.07) [68] |
| toxicity to various agents | 1,83 | 0.48 (0.07-0.77) [66] |
| gastrooesophageal reflux disease | 1,83 | 1.97 (1.57-2.27) [66] |
| fear | 1,83 | 2.1 (1.69-2.4) [66] |
| decreased appetite | 1,77 | 0.75 (0.34-1.05) [64] |
| hypoaesthesia | 1,69 | 1.1 (0.68-1.41) [61] |
| antipsychotic drug level below therapeutic | 1,66 | 3.69 (3.26-3.99) [60] |
| therapeutic product effect variable | 1,63 | 4.06 (3.63-4.37) [59] |
| hypersomnia | 1,61 | 1.61 (1.17-1.92) [58] |
| leukopenia | 1,55 | 1.79 (1.35-2.11) [56] |
| drug dependence | 1,55 | 1.88 (1.44-2.2) [56] |
| nightmare | 1,50 | 1.87 (1.42-2.19) [54] |
| unintended pregnancy | 1,50 | 3.71 (3.26-4.04) [54] |
| blood triglycerides increased | 1,47 | 1.15 (0.69-1.48) [53] |
| cough | 1,44 | 1.06 (0.6-1.39) [52] |
| poor quality sleep | 1,36 | 2.25 (1.78-2.59) [49] |
| abdominal pain | 1,36 | 0.74 (0.27-1.08) [49] |
| psychomotor hyperactivity | 1,33 | 1.32 (0.84-1.66) [48] |
| paranoia | 1,30 | 0.64 (0.15-0.98) [47] |
| gastrointestinal surgery | 1,30 | 4.06 (3.57-4.41) [47] |
| migraine | 1,27 | 1.2 (0.71-1.55) [46] |
| bipolar disorder | 1,27 | 0.96 (0.47-1.31) [46] |
| mood swings | 1,27 | 1.23 (0.74-1.58) [46] |
| dyslipidaemia | 1,25 | 3.12 (2.62-3.47) [45] |
| amnesia | 1,25 | 0.8 (0.31-1.16) [45] |
| thinking abnormal | 1,19 | 0.86 (0.35-1.22) [43] |
| affective disorder | 1,19 | 1.96 (1.45-2.32) [43] |
| dyspepsia | 1,16 | 1.8 (1.28-2.16) [42] |
| hunger | 1,16 | 2.14 (1.63-2.51) [42] |
| sleep apnoea syndrome | 1,16 | 1.3 (0.79-1.67) [42] |
| dependence | 1,16 | 3.29 (2.78-3.66) [42] |
| myalgia | 1,14 | 0.55 (0.03-0.92) [41] |
| crying | 1,14 | 0.81 (0.29-1.18) [41] |
| panic attack | 1,14 | 1.03 (0.51-1.4) [41] |
| incorrect dose administered | 1,14 | 0.69 (0.17-1.06) [41] |
| muscle twitching | 1,11 | 0.79 (0.26-1.16) [40] |
| prescribed underdose | 1,11 | 0.89 (0.37-1.27) [40] |
| personality disorder | 1,08 | 2.66 (2.13-3.04) [39] |
| apathy | 1,08 | 1.25 (0.71-1.63) [39] |
| alanine aminotransferase increased | 1,08 | 1.05 (0.52-1.43) [39] |
| loss of personal independence in daily activities | 1,05 | 1.12 (0.58-1.51) [38] |
| cognitive disorder | 1,03 | 0.79 (0.25-1.18) [37] |
| low density lipoprotein increased | 1,03 | 3.22 (2.67-3.61) [37] |

**Table S6. Events disproportionally reported with pramipexole-induced impulsivity**. Ordered by N.

| Event | Perc (%) | Information Component Median (95%CI) [N] |
|---|---|---|
| obsessive-compulsive disorder | 26,77 | 3.47 (3.33-3.57) [553] |
| emotional distress | 21,35 | 3.42 (3.26-3.54) [441] |
| Depression | 20,67 | 2.43 (2.27-2.55) [427] |
| Pain | 16,02 | 1.64 (1.45-1.77) [331] |
| Anxiety | 13,79 | 1.93 (1.73-2.07) [285] |
| suicidal ideation | 9,73 | 2.7 (2.47-2.87) [201] |
| weight increased | 9,29 | 1.93 (1.7-2.11) [192] |
| Insomnia | 7,12 | 0.79 (0.52-0.99) [147] |
| abnormal behaviour | 6,73 | 2.52 (2.24-2.72) [139] |
| Somnolence | 6,39 | 0.72 (0.43-0.92) [132] |
| economic problem | 6,05 | 3.15 (2.85-3.36) [125] |
| Stress | 5,86 | 2.5 (2.2-2.71) [121] |
| suicide attempt | 5,28 | 2.74 (2.43-2.97) [109] |
| Dyskinesia | 4,70 | 0.62 (0.28-0.86) [97] |
| Fear | 4,65 | 2.95 (2.61-3.19) [96] |
| Injury | 4,11 | 2.53 (2.17-2.78) [85] |
| Aggression | 3,05 | 1.9 (1.48-2.2) [63] |
| personality change | 2,66 | 2.93 (2.49-3.26) [55] |
| eating disorder | 2,47 | 2.95 (2.49-3.28) [51] |
| Dependence | 2,37 | 3.26 (2.79-3.6) [49] |
| hallucination, visual | 2,37 | 0.69 (0.21-1.03) [49] |
| psychomotor hyperactivity | 2,37 | 2.24 (1.76-2.58) [49] |
| dopamine dysregulation syndrome | 2,32 | 2.72 (2.24-3.07) [48] |
| Overdose | 2,27 | 1.27 (0.79-1.62) [47] |
| sleep disorder | 2,23 | 1.07 (0.58-1.42) [46] |
| psychotic disorder | 2,23 | 1.56 (1.06-1.91) [46] |
| Agitation | 2,18 | 0.81 (0.32-1.17) [45] |
| mental disorder | 2,13 | 2.24 (1.74-2.6) [44] |
| drug withdrawal syndrome | 1,94 | 1.78 (1.26-2.16) [40] |
| drug dependence | 1,94 | 2.34 (1.81-2.71) [40] |
| irritability | 1,89 | 1.95 (1.42-2.33) [39] |
| drug abuse | 1,89 | 2.46 (1.93-2.84) [39] |
| mania | 1,84 | 2.02 (1.48-2.4) [38] |
| on and off phenomenon | 1,79 | 0.87 (0.32-1.26) [37] |
| cognitive disorder | 1,74 | 1.04 (0.48-1.43) [36] |
| compulsions | 1,74 | 3.05 (2.49-3.44) [36] |
| sudden onset of sleep | 1,74 | 1.61 (1.05-2) [36] |
| crying | 1,60 | 1.86 (1.28-2.27) [33] |
| delusion | 1,60 | 0.93 (0.35-1.34) [33] |
| apathy | 1,45 | 2.7 (2.09-3.13) [30] |
| jealous delusion | 1,45 | 2.58 (1.98-3.02) [30] |
| restlessness | 1,40 | 0.69 (0.07-1.13) [29] |
| disturbance in attention | 1,40 | 1 (0.38-1.44) [29] |
| hypomania | 1,40 | 2.6 (1.98-3.04) [29] |
| bipolar disorder | 1,36 | 2.2 (1.57-2.64) [28] |
| thinking abnormal | 1,36 | 1.91 (1.28-2.36) [28] |
| withdrawal syndrome | 1,31 | 1.35 (0.71-1.8) [27] |
| depressed mood | 1,31 | 0.95 (0.31-1.4) [27] |
| major depression | 1,26 | 2.69 (2.03-3.15) [26] |
| anger | 1,21 | 2.04 (1.37-2.51) [25] |
| paranoia | 1,21 | 1.54 (0.87-2.01) [25] |
| marital problem | 1,11 | 3.3 (2.61-3.79) [23] |
| anhedonia | 1,11 | 2.09 (1.39-2.58) [23] |
| product use in unapproved indication | 1,11 | 0.78 (0.09-1.27) [23] |
| emotional disorder | 1,06 | 1.48 (0.76-1.98) [22] |
| mental impairment | 1,02 | 1.52 (0.79-2.03) [21] |

**Table S7. Nodes with highest degree-centrality.**

| Aripiprazole | | | | | |
|---|---|---|---|---|---|
| Ising | | Phi | | PPMI | |
| economic problem | 24 | irritability | 49 | overwork | 97 |
| gambling disorder | 20 | anxiety | 47 | pyromania | 89 |
| mental disorder | 16 | product use in unapproved indication | 44 | kleptomania | 85 |
| obsessive-compulsive disorder | 16 | suicide attempt | 41 | poriomania | 60 |
| compulsive shopping | 15 | obsessive-compulsive disorder | 39 | anxiety | 45 |
| Fatigue | 15 | depression | 38 | product use in unapproved indication | 43 |
| depressed mood | 15 | economic problem | 38 | suicide attempt | 39 |
| Insomnia | 15 | insomnia | 35 | obsessive-compulsive disorder | 39 |
| impulsivity | 14 | suicidal ideation | 35 | economic problem | 38 |
| emotional distress | 13 | asthenia | 34 | gambling disorder | 37 |

| Pramipexole | | | | | |
|---|---|---|---|---|---|
| Ising | | Phi | | PPMI | |
| gambling disorder | 10 | mental disorder | 20 | pyromania | 60 |
| emotional distress | 9 | agitation | 18 | poriomania | 57 |
| Dyskinesia | 7 | anxiety | 14 | gaming disorder | 55 |
| Anxiety | 7 | bipolar disorder | 14 | excessive exercise | 48 |
| depression | 7 | cognitive disorder | 14 | kleptomania | 47 |
| emotional disorder | 6 | crying | 14 | body-focused disorder | 39 |
| stereotypy | 6 | depression | 14 | paranoia | 32 |
| mental disorder | 5 | insomnia | 14 | emotional disorder | 31 |
| cognitive disorder | 5 | apathy | 13 | mental impairment | 31 |
| drug withdrawal syndrome | 5 | abnormal behaviour | 12 | product use in unapproved indication | 30 |

**Table S8. Aripiprazole edges.** The ten strongest links for the three networks of aripiprazole are shown. Note that the links are undirected, therefore the two nodes may be switched.

| Ising | | | φ | | | PPMI | | |
|---|---|---|---|---|---|---|---|---|
| antipsychotic drug level below therapeutic | 8.21 | therapeutic product effect variable | antipsychotic drug level below therapeutic | 0.99 | therapeutic product effect variable | overwork | 8.84 | pyromania |
| theft | 7.26 | shoplifting | antipsychotic drug level below therapeutic | 0.95 | toxicity | kleptomania | 8.33 | overwork |
| therapeutic product effect incomplete | 6.18 | therapeutic product effect variable | therapeutic product effect variable | 0.94 | toxicity | overwork | 7.08 | poriomania |
| toxicity to various agents | 6 | antipsychotic drug level below therapeutic | antipsychotic drug level below therapeutic | 0.93 | leukopenia | kleptomania | 6.96 | pyromania |
| mental impairment | 4.96 | social problem | leukopenia | 0.92 | therapeutic product effect variable | antipsychotic drug level below therapeutic | 5.95 | therapeutic product effect variable |
| obesity | 4.32 | dyslipidaemia | shoplifting | 0.91 | theft | antipsychotic drug level below therapeutic | 5.89 | leukopenia |
| therapeutic product effect incomplete | 4.16 | toxicity to various agents | leukopenia | 0.89 | toxicity | leukopenia | 5.89 | therapeutic product effect variable |
| suicide attempt | 4.16 | suicidal ideation | therapeutic product effect incomplete | 0.83 | therapeutic product effect variable | muscle twitching | 5.84 | personality disorder |
| euphoric mood | 4.1 | antipsychotic drug level below therapeutic | therapeutic product effect incomplete | 0.82 | toxicity | antipsychotic drug level below therapeutic | 5.81 | toxicity to various agents |
| gastrooesophageal reflux disease | 3.99 | migraine | antipsychotic drug level below therapeutic | 0.82 | therapeutic product effect incomplete | therapeutic product effect variable | 5.81 | toxicity to various agents |

**Table S9. Pramipexole edges.** The ten strongest links for the three networks of pramipexole are shown. Note that the links are undirected, therefore the two nodes may be switched.

| | Ising | | | φ | | | PPMI | |
|---|---|---|---|---|---|---|---|---|
| body-focused disorder | 3.11 | kleptomania | emotional distress | 0.56 | pain | poriomania | 8.82 | pyromania |
| mental impairment | 3.09 | mental disorder | emotional distress | 0.55 | OCD | gaming disorder | 8.39 | pyromania |
| on and off phenomenon | 3.05 | dyskinesia | hyperphagia | 0.5 | weight increased | gaming disorder | 7.8 | poriomania |
| hyperphagia | 2.67 | weight increased | anxiety | 0.48 | fear | excessive exercise | 7.32 | pyromania |
| paranoia | 2.66 | psychotic disorder | anxiety | 0.47 | stress | kleptomania | 7.32 | pyromania |
| anxiety | 2.61 | fear | anxiety | 0.43 | depression | excessive exercise | 6.74 | poriomania |
| jealous delusion | 2.46 | marital problem | depression | 0.41 | stress | kleptomania | 6.74 | poriomania |
| pain | 2.36 | emotional distress | fear | 0.41 | pain | paranoia | 6.39 | pyromania |
| cognitive disorder | 2.36 | emotional disorder | mental disorder | 0.39 | mental impairment | excessive exercise | 3.3 | gaming disorder |
| depressed mood | 2.3 | anhedonia | dyskinesia | 0.38 | on and off phenomenon | gaming disorder | 3.3 | kleptomania |

**Table S10. Aripiprazole clusters obtained from Ising and Phi estimation.** Phi and Ising clusters are shown side by side. Ising clusters are color-coded, to show the similiarities with Phi.

| Ising | PHI | PPMI |
|---|---|---|
| Hypersomnia; sleep disorder; insomnia; somnolence | Hypersomnia; sleep disorder; insomnia | Hypersomnia; sleep disorder; insomnia |
| Fatigue; asthenia; feeling abnormal | Fatigue; asthenia; feeling abnormal | Fatigue; asthenia; feeling abnormal |
| Constipation | Constipation; abdominal pain | Constipation; abdominal pain |
| Decreased appetite; weight decreased | Decreased appetite | Decreased appetite |
| Panic attack; stress; bipolar disorder; crying; depressed mood; depression; apathy; mood swings; mania | Panick attack; stress; bipolar disorder; crying; depressed mood; depression; apathy; mood swings; mania; affective disorder | Panick attack; stress; bipolar disorder; crying; depressed mood; depression; apathy; mood swings; mania; affective disorder |
| Aggression; anger; irritability | Aggression; anger | Aggression; anger |
| Psychomotor hyperactivity; restlessness; agitation | Psychomotor hyperactivity; restlessness; agitation | Psychomotor hyperactivity; restlessness; agitation |
| Abnormal behaviour | Abnormal behaviour; thinking abnormal | Abnormal behaviour; thinking abnormal |
| Myalgia | Myalgia; arthralgia; back pain | Myalgia; arthralgia; back pain |
| Drug dependence | Drug dependence; dependence | Drug dependence; dependence |
| Disturbance in attention; memory impairment | Disturbance in attention; amnesia; cognitive disorder | Disturbance in attention; amnesia; cognitive disorder |
| Hallucination, auditory | Hallucination, auditory; paranoia | Hallucination, auditory; paranoia |
| **impulsivity** Paraphilia; stereotypy | Paraphilia; stereotypy; kleptomania; poriomania | Paraphilia; stereotypy; kleptomania; poriomania; **overwork; pyromania** |
| Headache | Cough | Cough |
| | Loss of personal independence in daily activities | Loss of personal independence in daily activities |
| | Hospitalisation | Hospitalisation |
| | Prescribed underdose**; prescribed overdose** | Prescribed underdose; **incorrect dose administered** |
| Blood triglycerides increased; low density lipoprotein increased; blood cholesterol increased; blood glucose increased; alanine aminotransferase increased | Blood triglycerides increased; low density lipoprotein increased; blood cholesterol increased; blood glucose increased; alanine aminotransferase increased | Blood triglycerides increased; low density lipoprotein increased; blood cholesterol increased; blood glucose increased; alanine aminotransferase increased; **hunger;** sleep apnoea syndrome; **gastrointestinal surgery** |
| Fear; affective disorder | Fear | Fear |
| Personal relationship issue; social problem | Personal relationship issue; social problem; theft; shoplifting; Disability; economic problem; bankruptcy; divorced; homeless; loss of employment | Personal relationship issue; social problem; theft; shoplifting; Disability; economic problem; bankruptcy; divorced; homeless; loss of employment |
| Mental impairment | Mental impairment | Mental impairment |

| | | |
|---|---|---|
| Pain<br>Emotional distress; anhedonia; neuropsychiatric symptoms; anxiety<br>Mental disorder<br>Injury; brain injury<br>Disability; economic problem; bankruptcy; divorced; homeless; loss of employment<br>**impulsivity** Gambling disorder; compulsive shopping; body-focused disorder; compulsive hoarding; hypersexuality; impulsivity<br>Unintended pregnancy; sexual dysfunction; psychosexual disorder; sexually transmitted disease<br>Obsessive-compulsive disorder<br>Eating disorder; gastrointestinal surgery<br>Condition aggravated<br>Product used in unapproved indication | Pain<br>Emotional distress; anhedonia; neuropsychiatric symptoms; anxiety<br>Mental disorder<br>Injury; brain injury<br>Gambling disorder; compulsive shopping; body-focused disorder; compulsive hoarding; hypersexuality; **impulsivity**<br>Unintended pregnancy; sexual dysfunction; psychosexual disorder; sexually transmitted disease<br>**Obsessive-compulsive disorder**<br>Eating disorder; **gastrointestinal surgery**<br>Condition aggravated<br>Product used in unapproved indication | Pain<br>Emotional distress; anhedonia; neuropsychiatric symptoms; anxiety<br>Mental disorder<br>Injury; brain injury<br>Gambling disorder; compulsive shopping; body-focused disorder; compulsive hoarding; hypersexuality<br>Unintended pregnancy; sexual dysfunction; psychosexual disorder; sexually transmitted disease<br>Eating disorder<br>Condition aggravated<br>Product used in unapproved indication |
| Theft; shoplifting<br>Suicidal ideation; suicide attempt | Suicidal ideation; suicide attempt | Suicidal ideation |
| Therapeutic product effect incomplete; therapeutic product effect variable; antipsychotic drug level below therapeutic; toxicity to various agents<br>Leukopenia<br>Euphoric mood | Therapeutic product effect incomplete; therapeutic product effect variable; antipsychotic drug level below therapeutic; toxicity to various agents; prescribed overdose<br>Leukopenia<br>Euphoric mood | Therapeutic product effect incomplete; therapeutic product effect variable; antipsychotic drug level below therapeutic; toxicity to various agents; prescribed overdose<br>Leukopenia<br>Euphoric mood |
| Hunger; hyperphagia; weight increased | Hunger; hyperphagia; weight increased; obesity; dyslipidaemia; hypertension; **sleep apnoea syndrome**<br>Akathisia<br>**impulsivity** Impulsivity | Obesity; dyslipidaemia; hypertension<br>Akathisia<br>Impulsivity<br>Suicide attempt<br>**Obsessive-compulsive disorder** |
| Obesity; dyslipidaemia; hypertension; sleep apnoea syndrome<br>Akathisia<br>Sedation<br>Amnesia; cognitive disorder | | |

| | | |
|---|---|---|
| 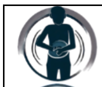 Diarrhoea; dyspepsia; gastroesophageal reflux disease; abdominal pain | Diarrhoea; dyspepsia; gastroesophageal reflux disease | Diarrhoea; dyspepsia; gastroesophageal reflux disease |
| 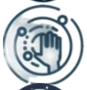 Hypoaesthesia | Hypoaesthesia | Hypoaesthesia |
| 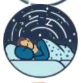 Poor quality sleep; nightmare | Poor quality sleep; nightmare; sedation | Poor quality sleep; nightmare; sedation; somnolence |
| 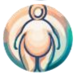 Hyperidrosis | Hyperidrosis | Hyperidrosis |
| 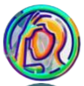 Migraine | Migraine; headache | Migraine; headache |
| 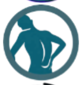 Muscle twitching; back pain; arthralgia | Muscle twitching | Muscle twitching |
| 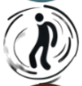 Personality disorder | Personality disorder | Personality disorder |
| 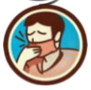 Cough | | |
| | 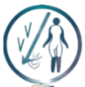 Weight decreased | Weight decreased |
| | 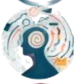 Memory impairment | Memory impairment |
| | 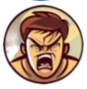 Irritability | Irritability |
| | | 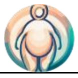 Hyperphagia; weight increased |
| **Isolated nodes**: pyromania; overwork; incorrect dose administered; prescribed overdose; prescribed underdose; poriomania; dependence; thinking abnormal; hospitalisation; paranoia; kleptomania; loss of personal independence in daily activities | **Isolated nodes**: pyromania; overwork; incorrect dose administered | |

**Table S11. Pramipexole clusters obtained from Ising and Phi estimation.** Phi and Ising clusters are shown side by side. Ising clusters are color-coded, to show the similiarities with Phi.

| Ising | PHI | PPMI |
|---|---|---|
| **impulsivity** Gambling disorder; compulsive shopping<br>Anxiety; depression; fear; emotional distress; stress<br>Pain<br>Obsessive-compulsive disorder<br>Suicidal ideation | Gambling disorder<br>Anxiety; depression; fear; emotional distress; stress; major depression<br>Pain<br>Obsessive-compulsive disorder<br>Suicidal ideation; suicide attempt<br>Injury | Gambling disorder; compulsive shopping<br>Depression; fear; emotional distress; stress<br>Pain<br>Obsessive-compulsive disorder<br>Suicidal ideation |
| Hyperphagia; weight increased | Hyperphagia; weight increased | Hyperphagia; weight increased |
| Sleep disorder; somnolence; insomnia; disturbance in attention | Sleep disorder; somnolence | |
| Drug withdrawal syndrome; on and off phenomenon; dyskinesia; dopamine dysregulation syndrome<br>**impulsivity** Stereotypy; impulsivity; paraphilia | Drug withdrawal syndrome; on and off phenomenon; dyskinesia; dopamine dysregulation syndrome; drug dependence; drug abuse<br>Stereotypy; impulsivity; paraphilia<br>Aggression<br>Compulsive hoarding<br>Paranoia; hallucination, visual; psychotic disorder<br>Psychomotor hyperactivity; restlessness<br>Hypomania; mania<br>Eating disorder | Drug withdrawal syndrome; on and off phenomenon; dyskinesia; dopamine dysregulation syndrome; drug dependence; drug abuse; dependence<br>Stereotypy; impulsivity; paraphilia; gaming disorder; poriomania; pyromania<br>Aggression<br>Compulsive hoarding<br>Hallucination, visual<br>Psychomotor hyperactivity; restlessness<br>Eating disorder<br>Economic problem; abnormal behaviour<br>Injury<br>Personality change |
| Drug dependence; drug abuse<br>Aggression<br>**impulsivity** Compulsive hoarding | | |

| | | |
|---|---|---|
| | | Somnolence<br>Suicide attempt |
| Bipolar disorder<br>Mental impairment; mental disorder<br>Abnormal behaviour<br>Economic problem<br>Product use in unapproved indication<br>Injury | Bipolar disorder; emotional disorder; apathy; crying<br>Mental impairment; mental disorder; cognitive disorder; disturbance in attention<br>Abnormal behaviour<br>Overdose<br>Agitation<br>Insomnia | Bipolar disorder; emotional disorder; apathy; crying; anxiety<br>Mental impairment; mental disorder; cognitive disorder; disturbance in attention<br>Overdose<br>Agitation<br>Insomnia |
| Anhedonia; depressed mood; emotional disorder; apathy<br>Cognitive disorder<br>Overdose<br>Suicide attempt<br>Delusion | | |
| Agitation<br>Crying<br>Irritability | | |
| Jealous delusion; paranoia; hallucination, visual; psychotic disorder<br>**impulsivity** Hypersexuality<br>Marital problem<br>Psychomotor hyperactivity<br>Hypomania; mania | Jealous delusion<br>Hypersexuality; excessive exercise; compulsive shopping<br>Marital problem;<br>Anhedonia; depressed mood | Jealous delusion; marital problem; paranoia; psychotic disorder; delusion<br>Hypersexuality; excessive exercise; body-focused disorder; kleptomania<br>Hypomania; mania; anhedonia; depressed mood; major depression |

| | | |
|---|---|---|
| | 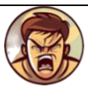 Irritability; anger<br>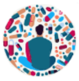 Dependence; withdrawal syndrome<br>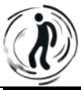 Thinking abnormal | Irritability; anger<br>Withdrawal syndrome<br>Thinking abnormal |
| **impulsivity** body-focused disorder; kleptomania | body-focused disorder; kleptomania | 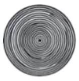 Compulsions<br>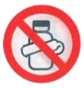 Product use in unapproved indication<br>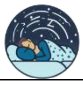 Sudden onset of sleep; sleep disorder |
| **Isolated nodes**: poriomania; compulsions; personality change; gaming disorder; eating disorder; sudden onset of sleep; restlessness; pyromania; thinking abnormal; withdrawal syndrome; excessive exercise; anger; dependence; major depression | **Isolated nodes**: poriomania; compulsions; personality change; gaming disorder | |

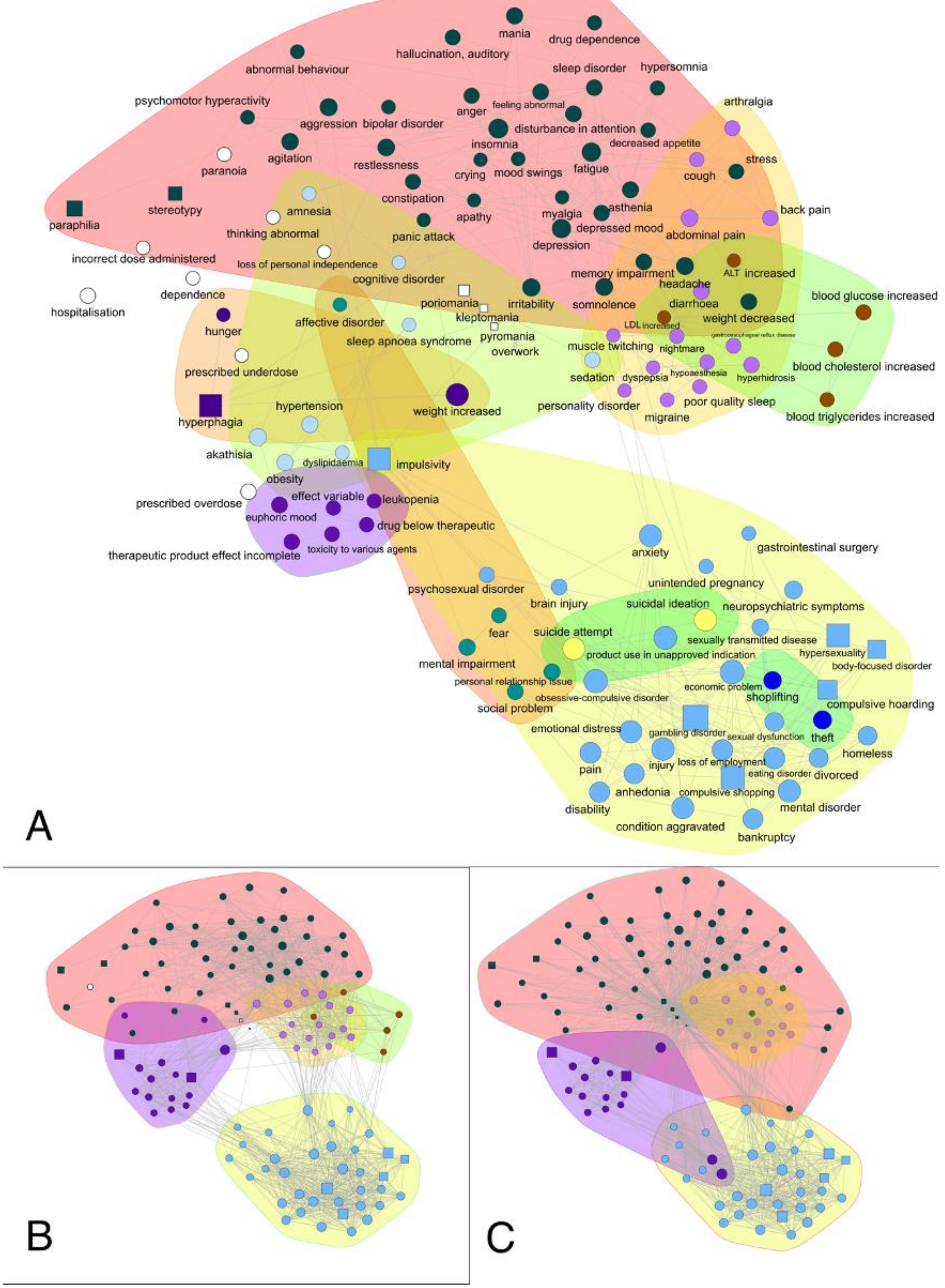


**Figure S1. The secondary impact of aripiprazole-induced pathologic impulsivity.** The network shows the events disproportionally reported with aripiprazole-related impulsivity. Impulsivity is shown as squares, other events as circles. Node colors and contours identify the clusters for the three methods: Ising (A), Phi (B) and PPMI (C). The layout is calculated using a spring model with, as weight, the average of the three weights calculated, after rescaling them from 0 to 1.

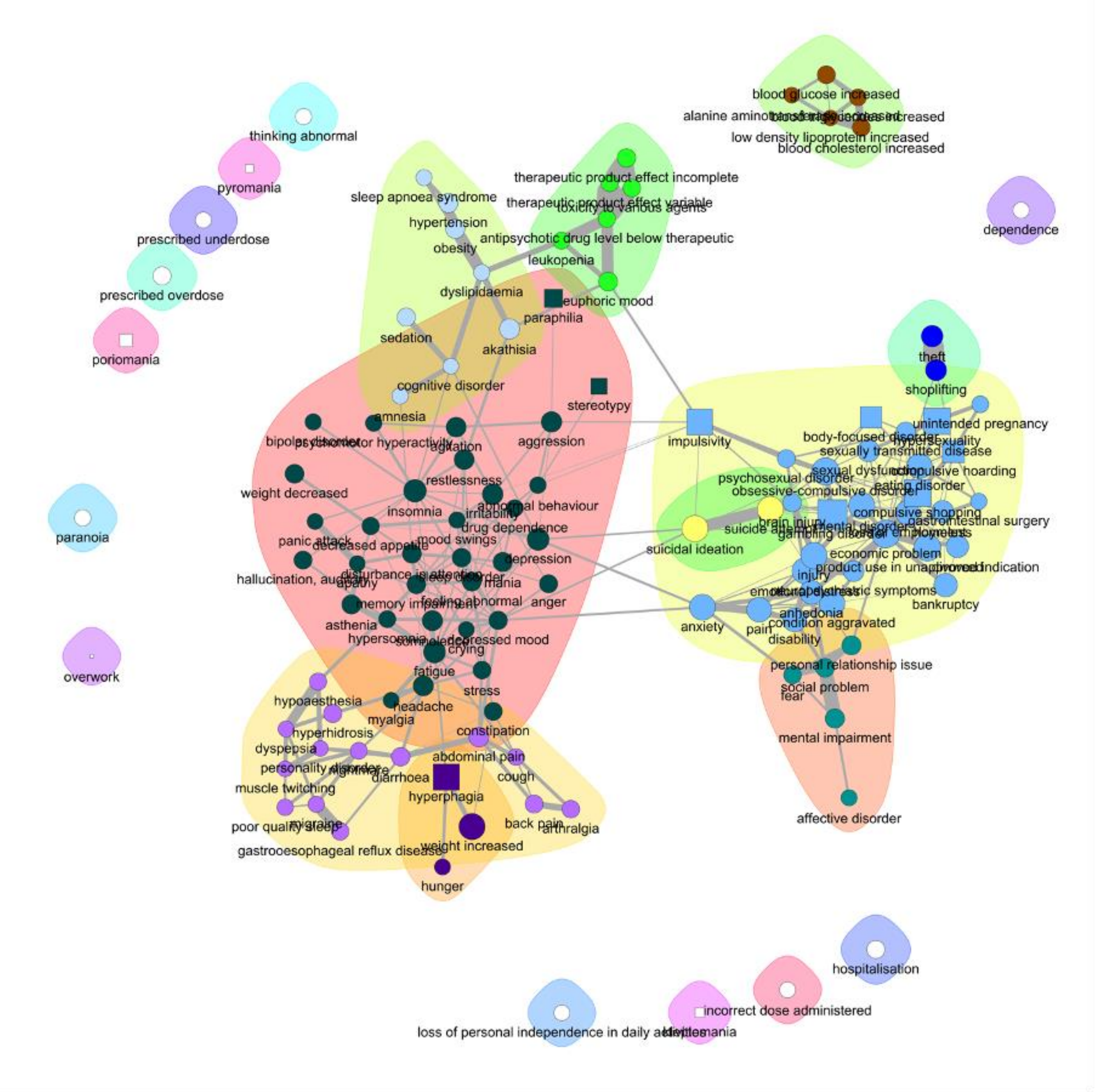


**Figure S2. The secondary impact of aripiprazole-induced pathologic impulsivity, Ising estimation.** The network shows the events disproportionally reported with aripiprazole-related impulsivity. Impulsivity is shown as squares, other events as circles. Node colors and contours identify the clusters for the Ising estimation. The layout is calculated using a spring model with, as weight, the links from the Ising.

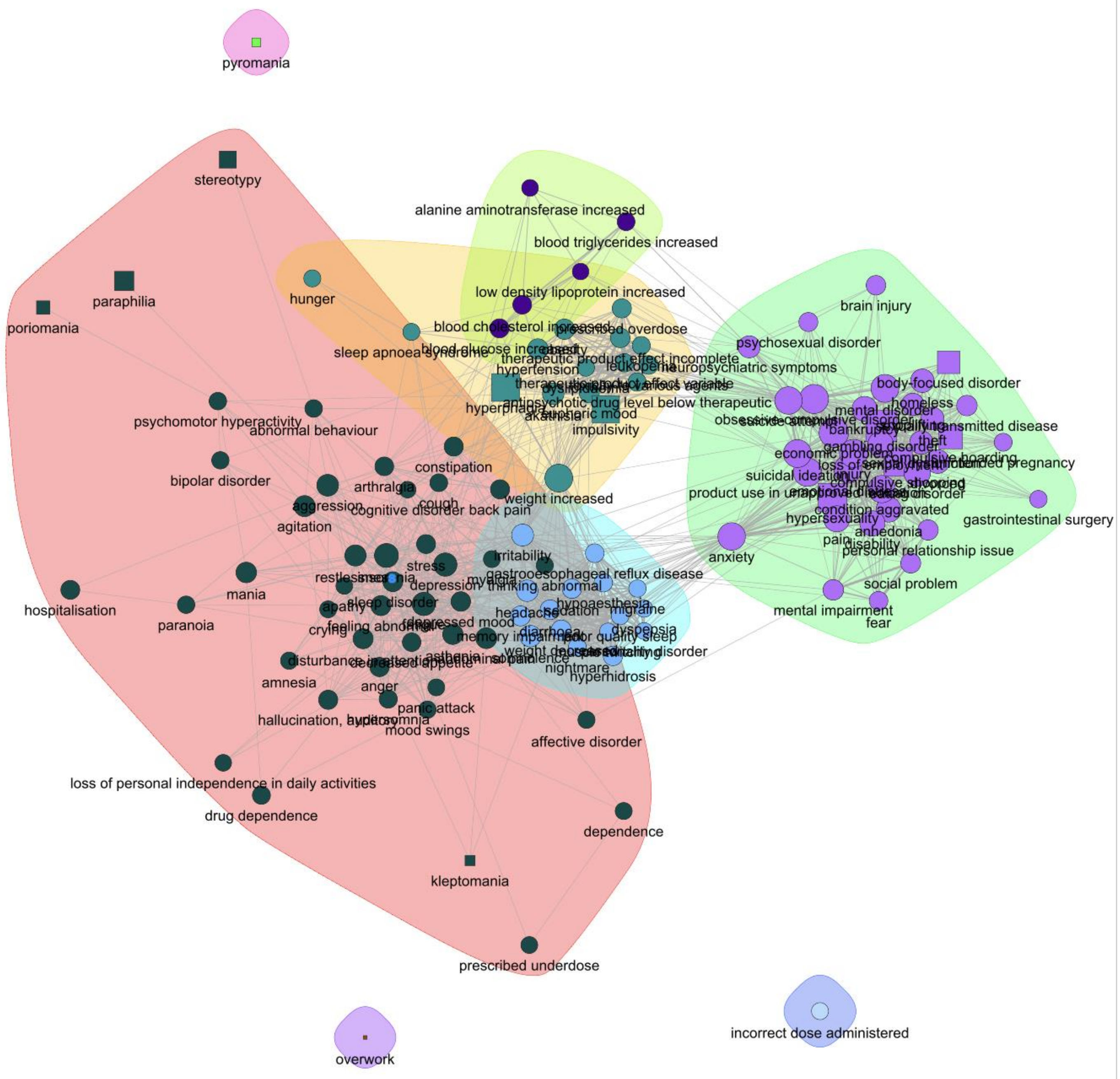


**Figure S3. The secondary impact of aripiprazole-induced pathologic impulsivity, Phi estimation.** The network shows the events disproportionally reported with aripiprazole-related impulsivity. Impulsivity is shown as squares, other events as circles. Node colors and contours identify the clusters for the Phi estimation. The layout is calculated using a spring model with, as weight, the links from the Phi.

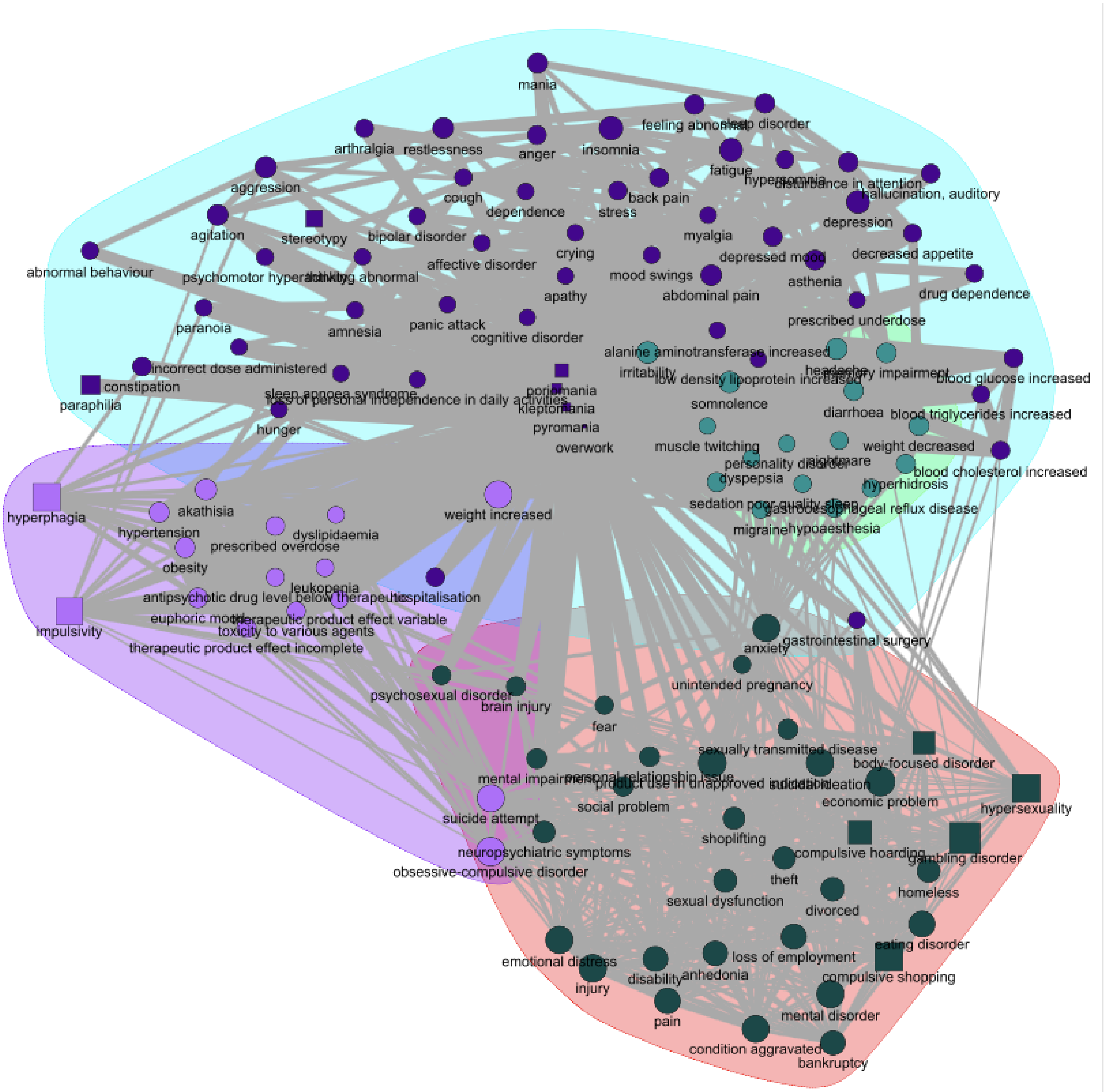


**Figure S4. The secondary impact of aripiprazole-induced pathologic impulsivity, PPMI estimation.** The network shows the events disproportionally reported with aripiprazole-related impulsivity. Impulsivity is shown as squares, other events as circles. Node colors and contours identify the clusters for the PPMI estimation. The layout is calculated using a spring model with, as weight, the links from the PPMI.

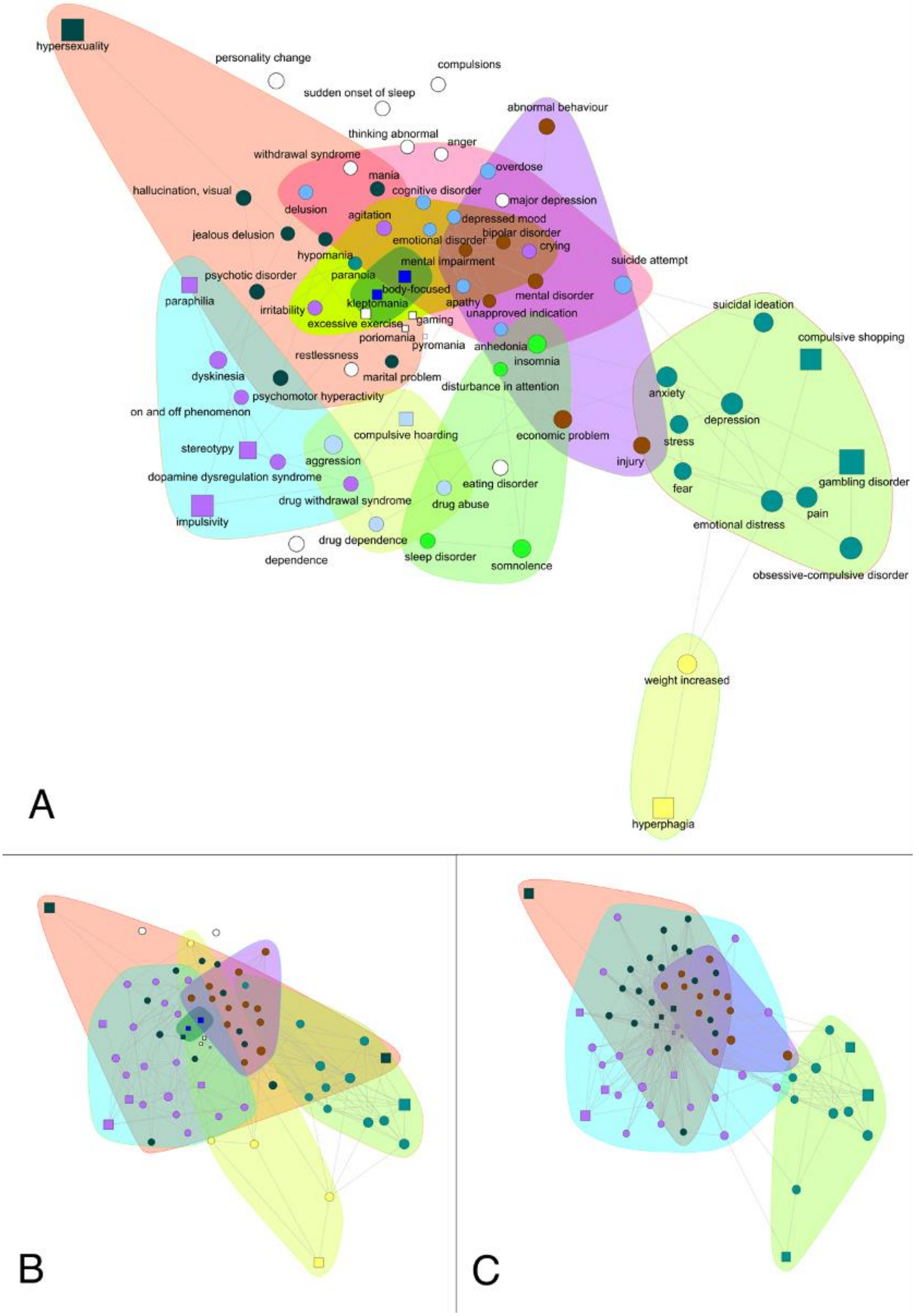


**Figure S5. The secondary impact of pramipexole-induced pathologic impulsivity.** The network shows the events disproportionally reported with pramipexole-related impulsivity. Impulsivity is shown as squares, other events as circles. Node colors and contours identify the clusters for the three methods: Ising (A), Phi (B) and PPMI (C). The layout is calculated using a spring model with, as weight, the average of the three weights calculated, after rescaling them from 0 to 1.

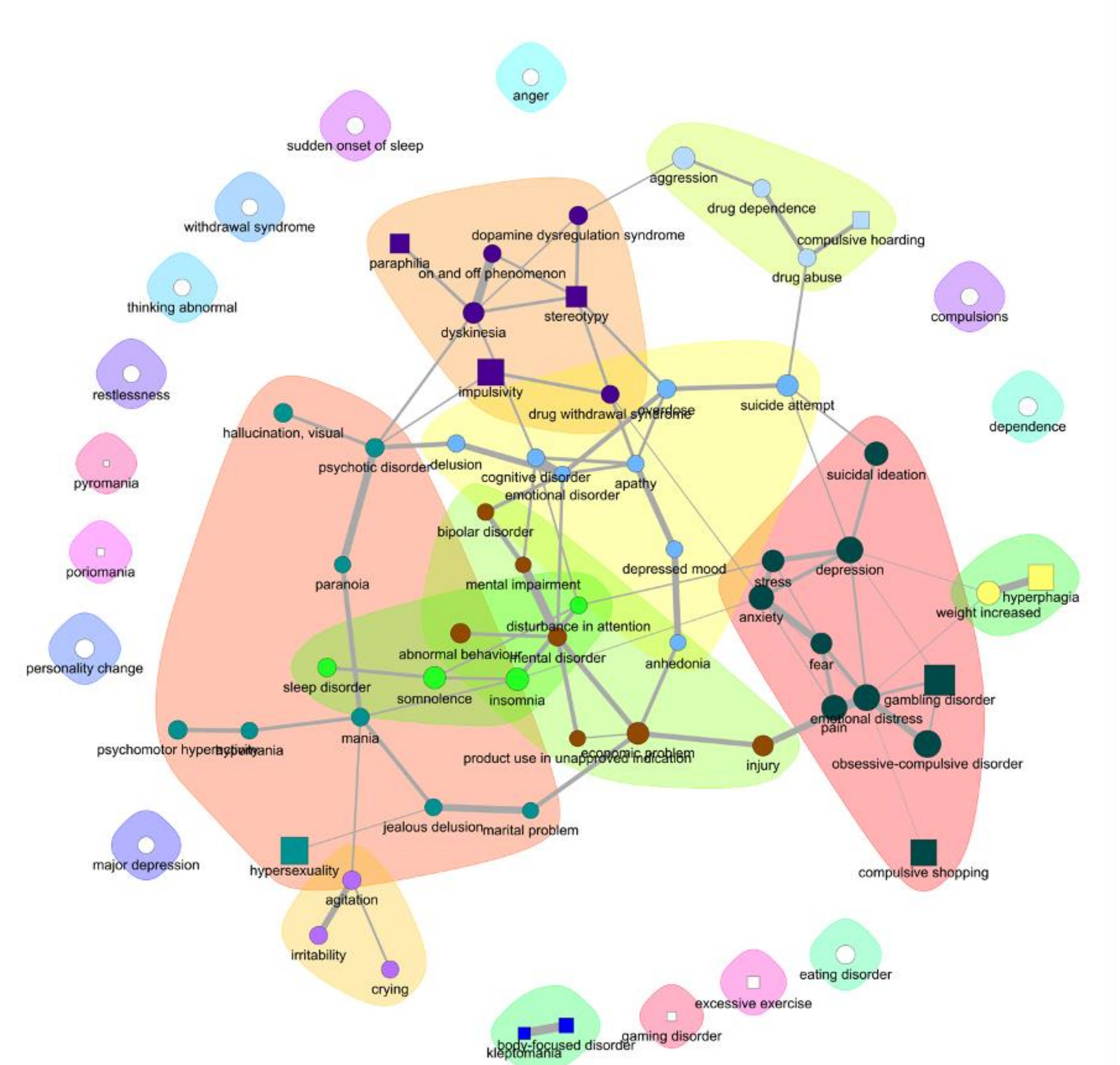


**Figure S6. The secondary impact of pramipexole-induced pathologic impulsivity, Ising estimation.** The network shows the events disproportionally reported with pramipexole-related impulsivity. Impulsivity is shown as squares, other events as circles. Node colors and contours identify the clusters for the Ising estimation. The layout is calculated using a spring model with, as weight, the links from the Ising.

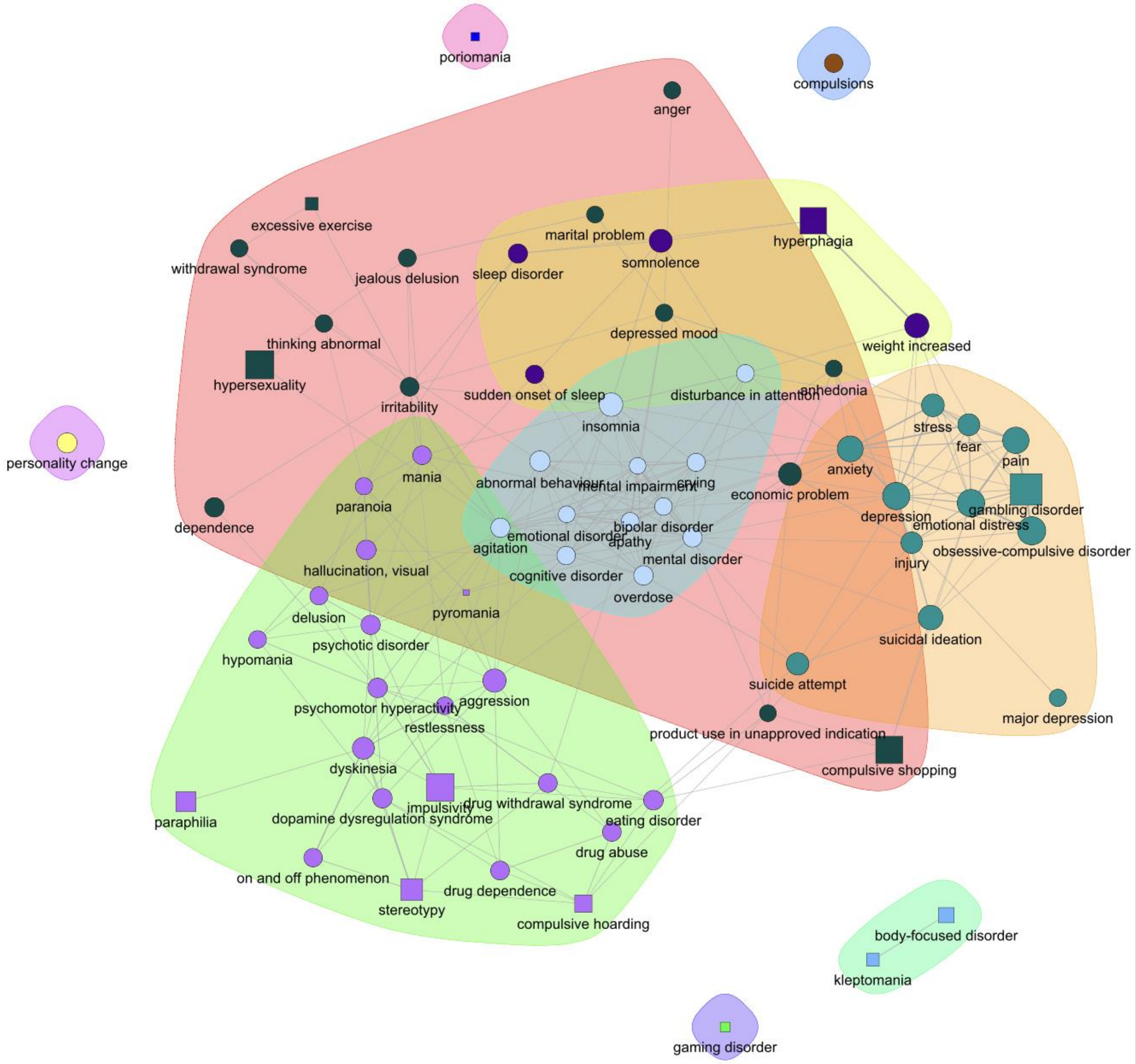


**Figure S7. The secondary impact of pramipexole-induced pathologic impulsivity, Phi estimation.** The network shows the events disproportionally reported with pramipexole-related impulsivity. Impulsivity is shown as squares, other events as circles. Node colors and contours identify the clusters for the Phi estimation. The layout is calculated using a spring model with, as weight, the links from the Phi.

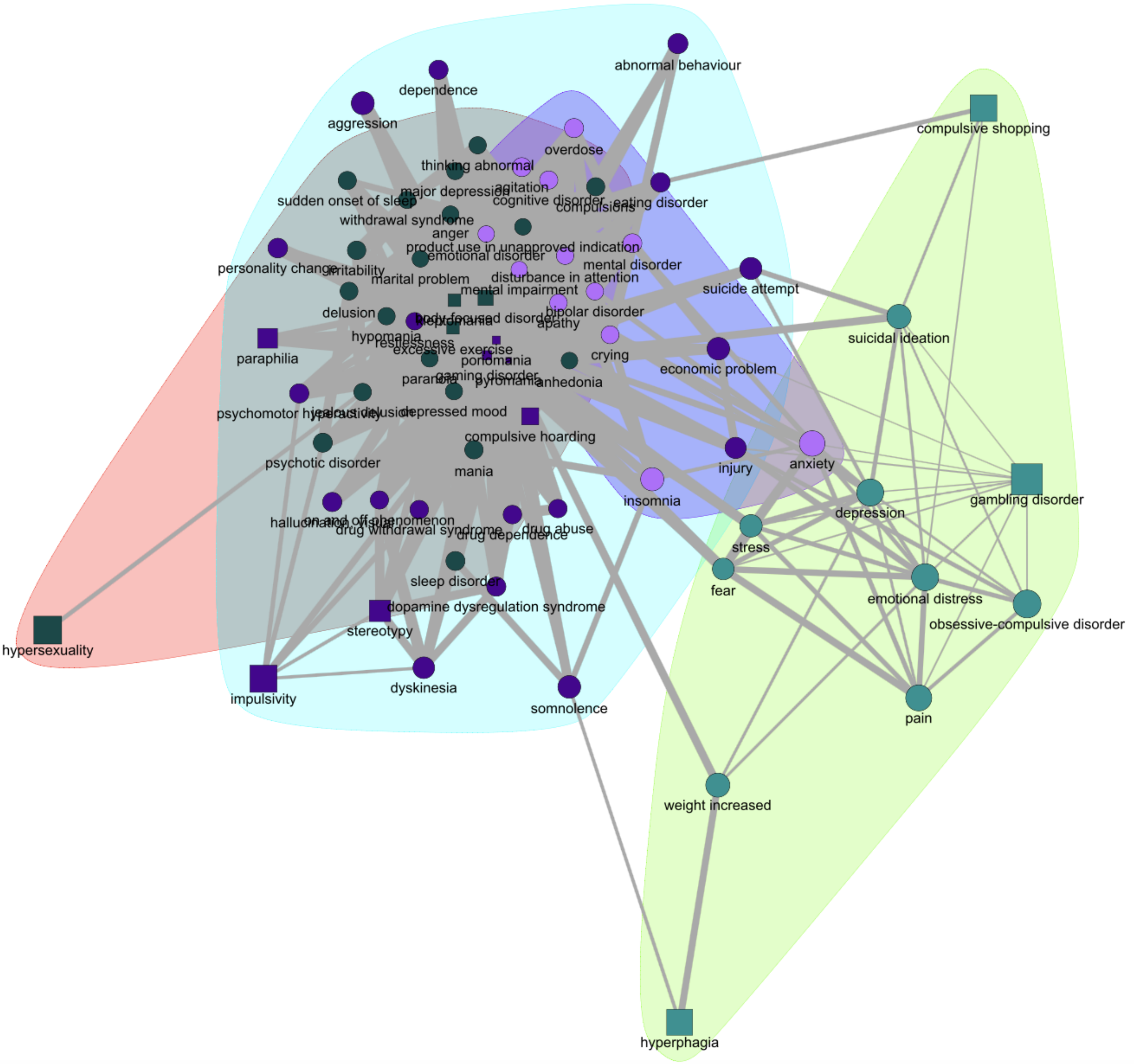


**Figure S8. The secondary impact of pramipexole-induced pathologic impulsivity, PPMI estimation.** The network shows the events disproportionally reported with pramipexole-related impulsivity. Impulsivity is shown as squares, other events as circles. Node colors and contours identify the clusters for the PPMI estimation. The layout is calculated using a spring model with, as weight, the links from the PPMI.

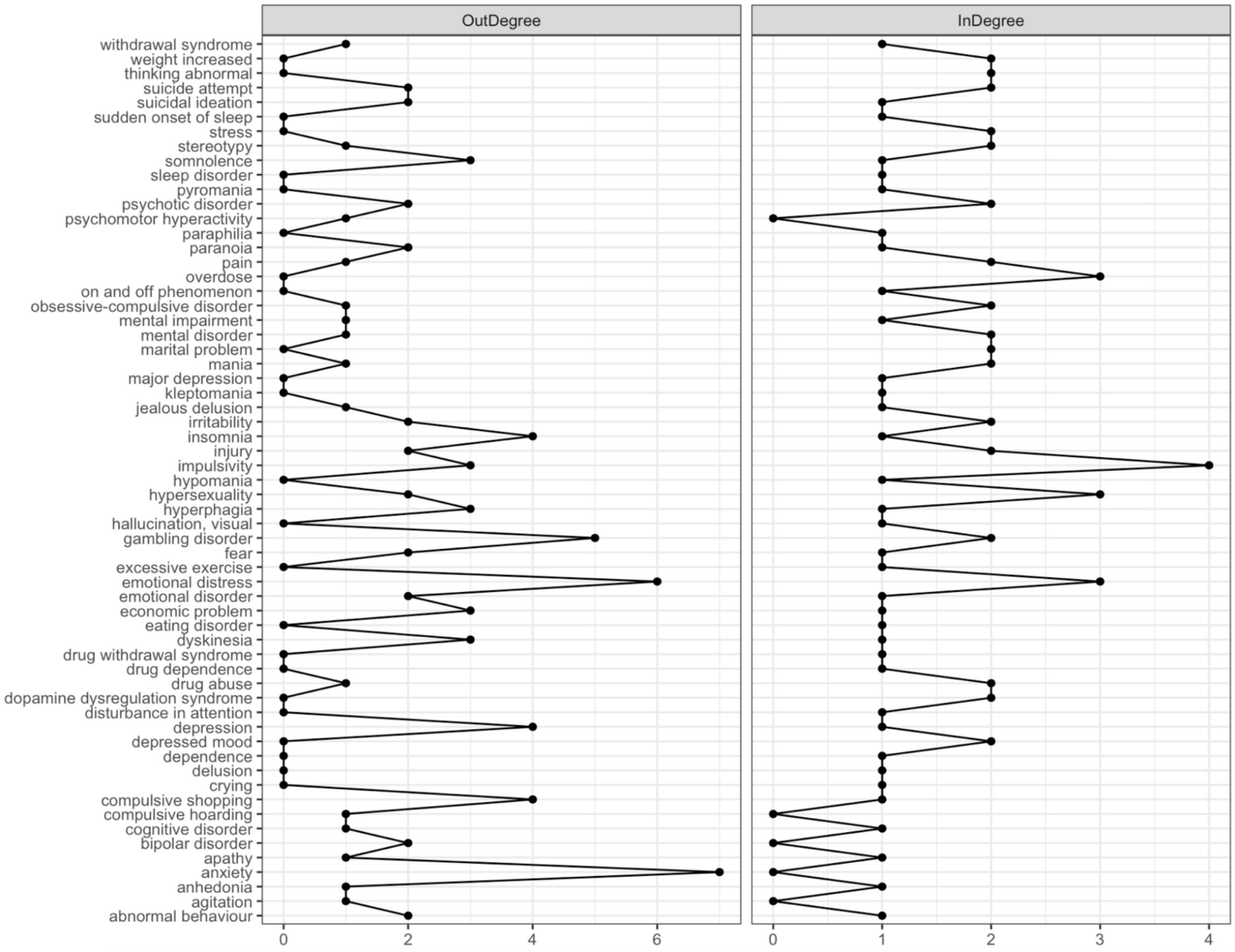


**Figure S9. Degree centrality, pramipexole.**

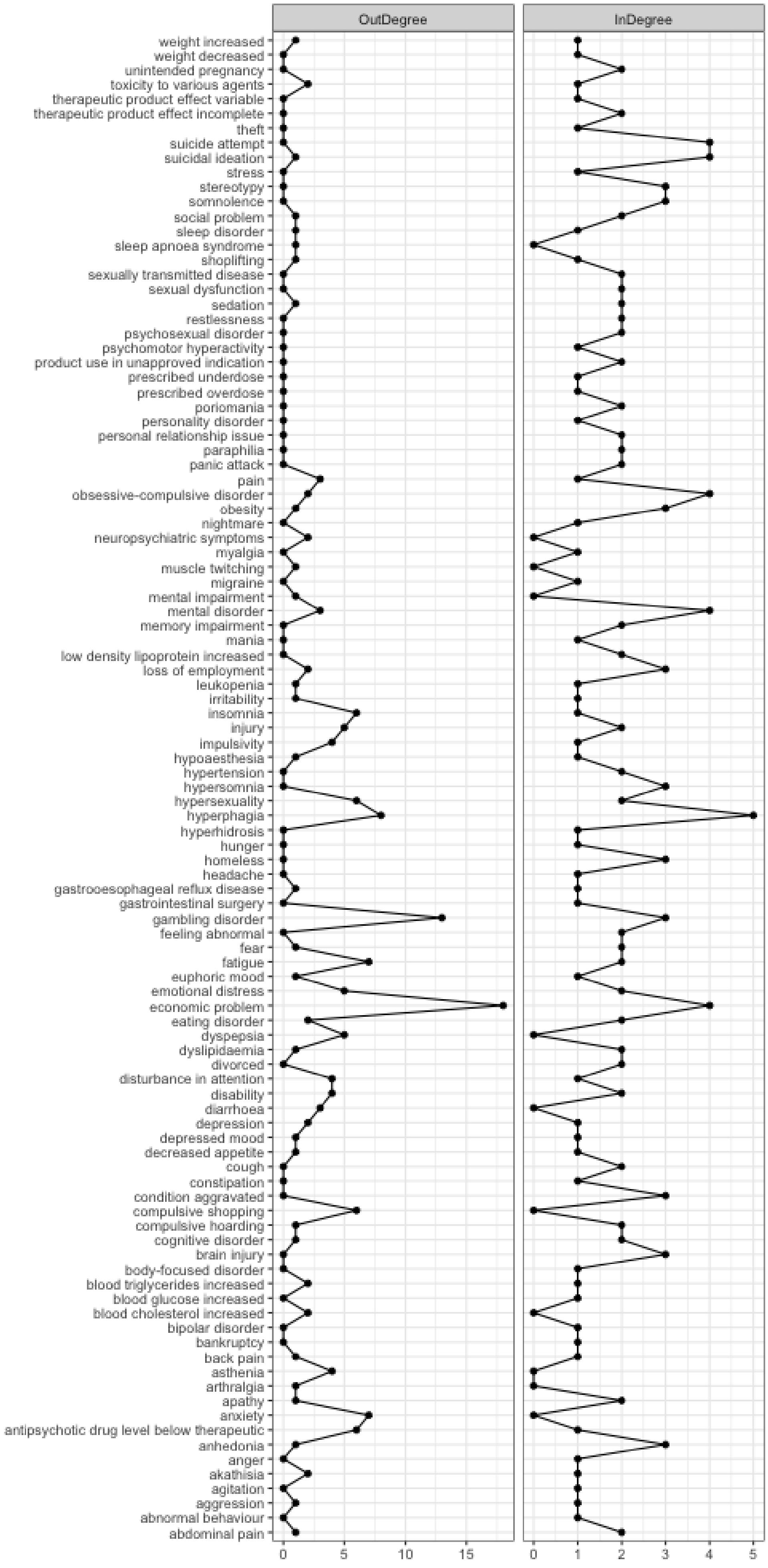


**Figure S10. Degree centrality, aripiprazole.**

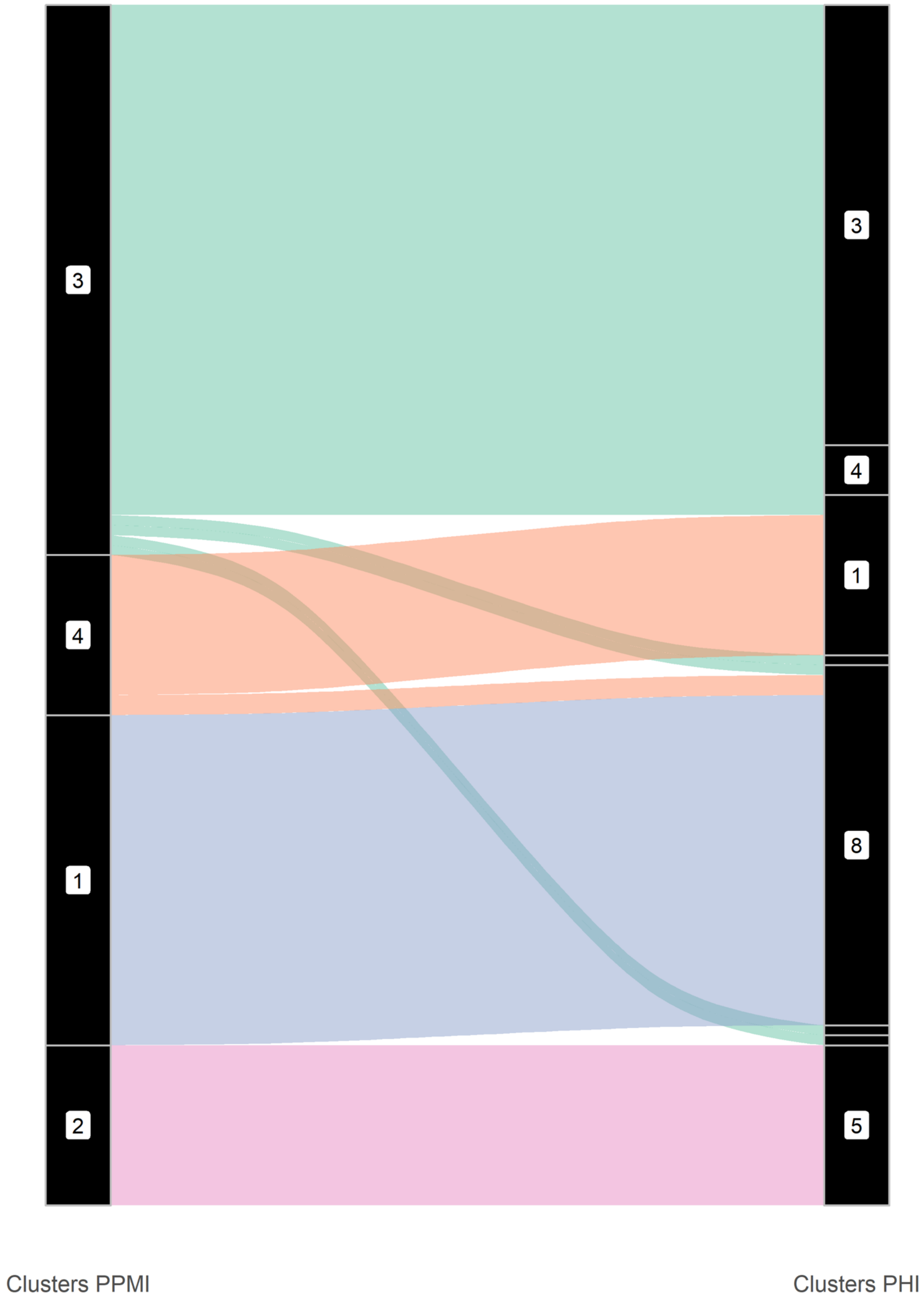


**Figure S11. Sankey showing comparison between clusters in PPMI and PHI, pramipexole.**

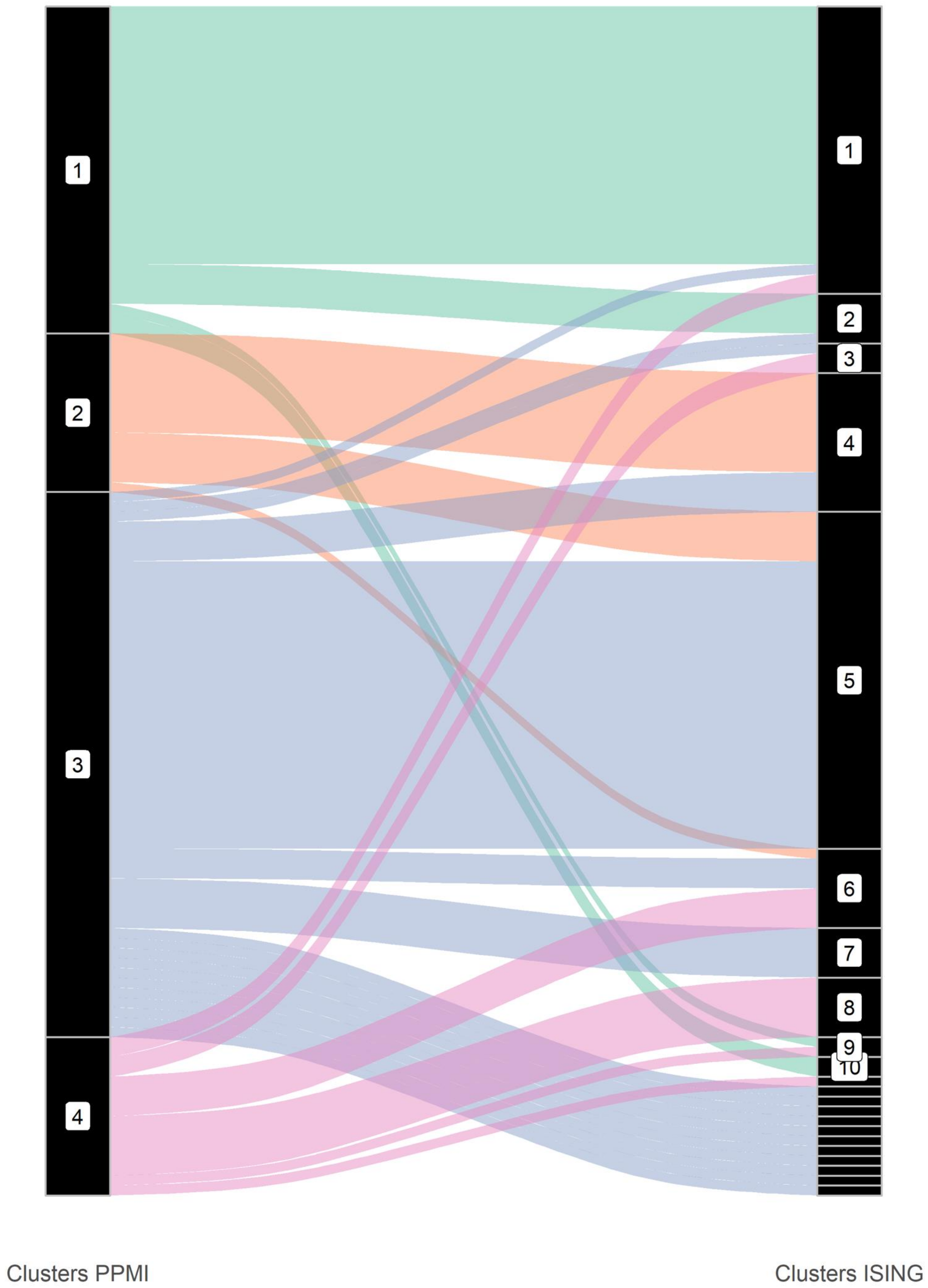


**Figure S12. Sankey showing comparison between clusters in PPMI and Ising, pramipexole.**

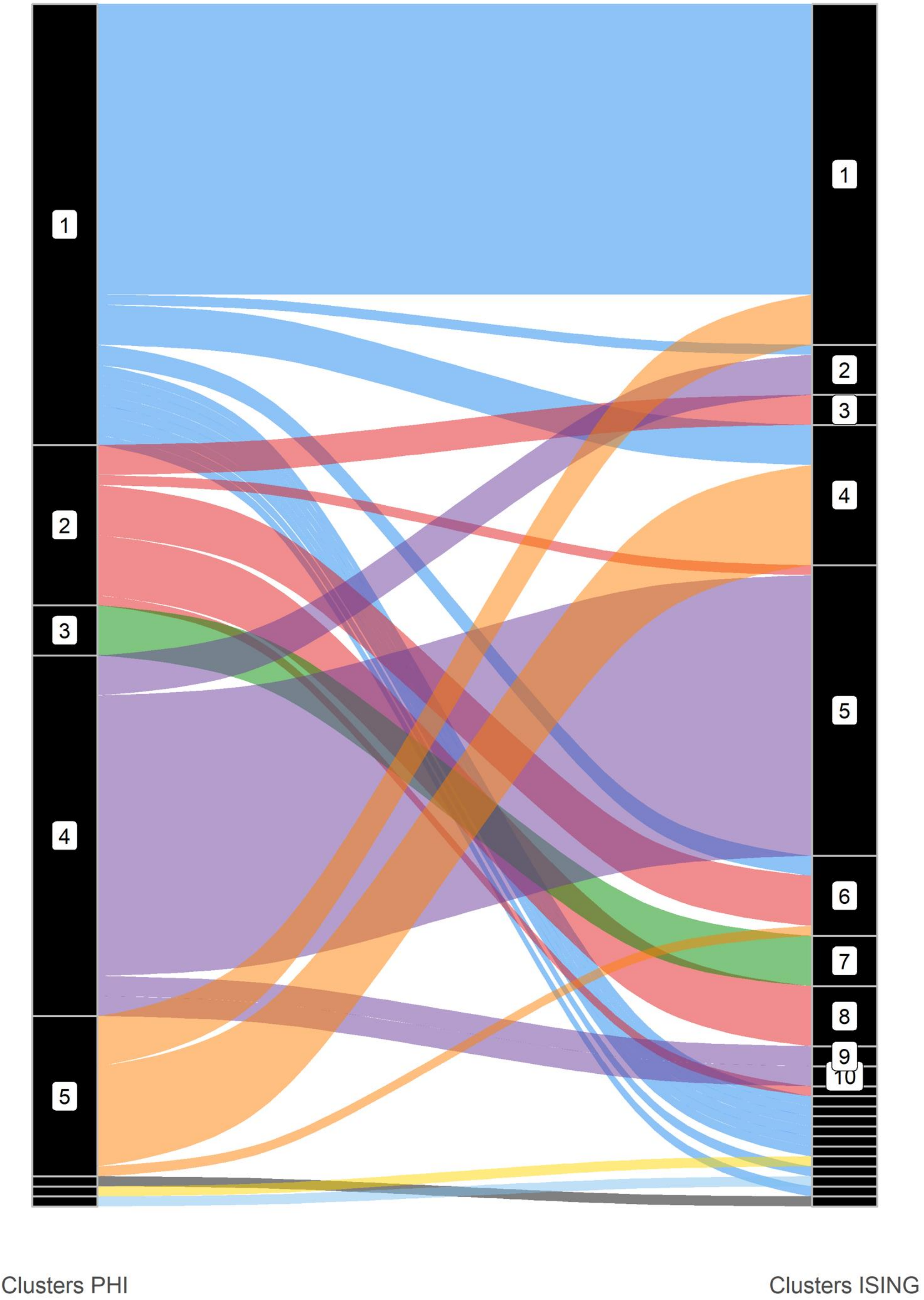


**Figure S13. Sankey showing comparison between clusters in Phi and Ising, pramipexole.**

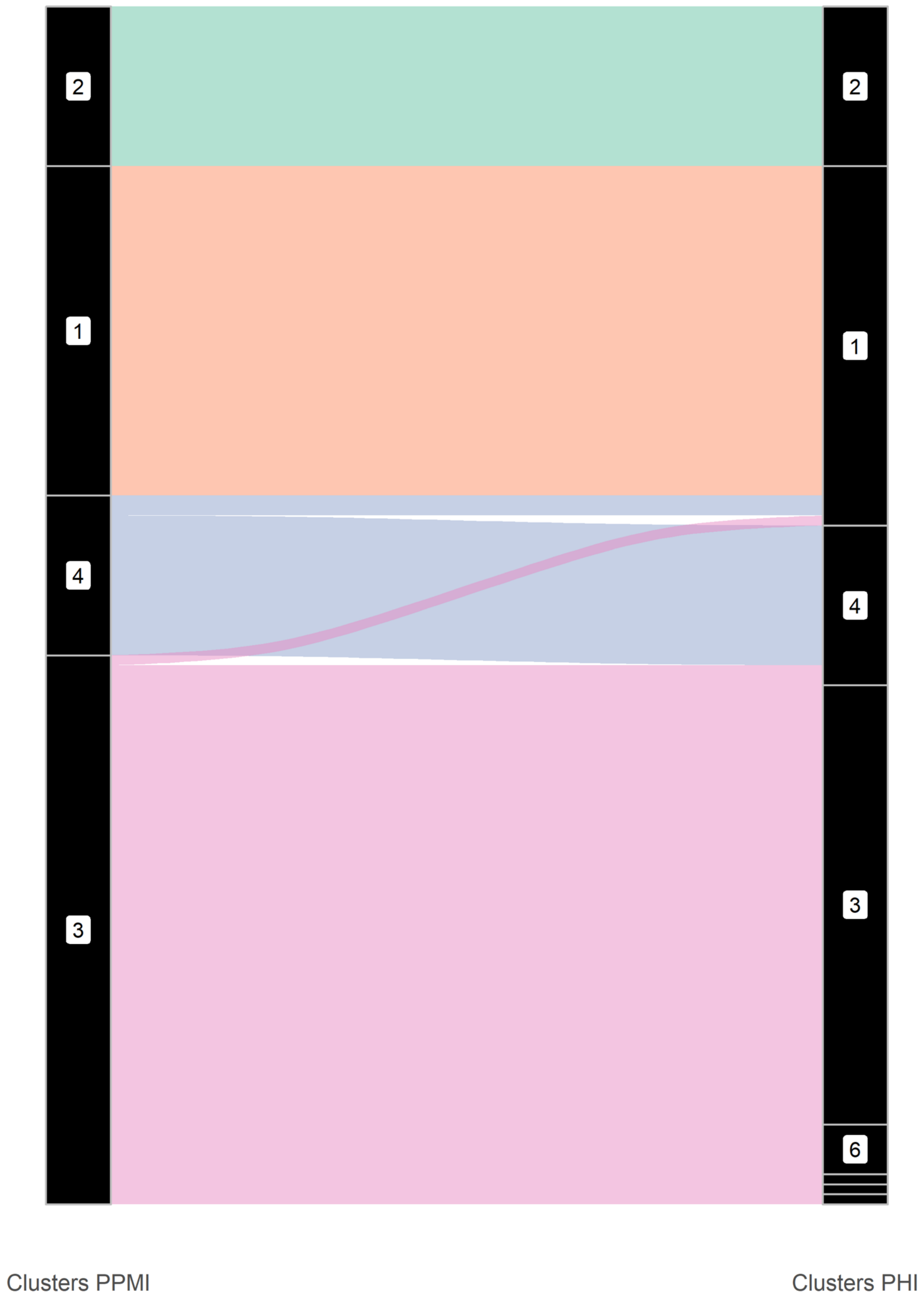


**Figure S14. Sankey showing comparison between clusters in PPMI and PHI, aripiprazole.**

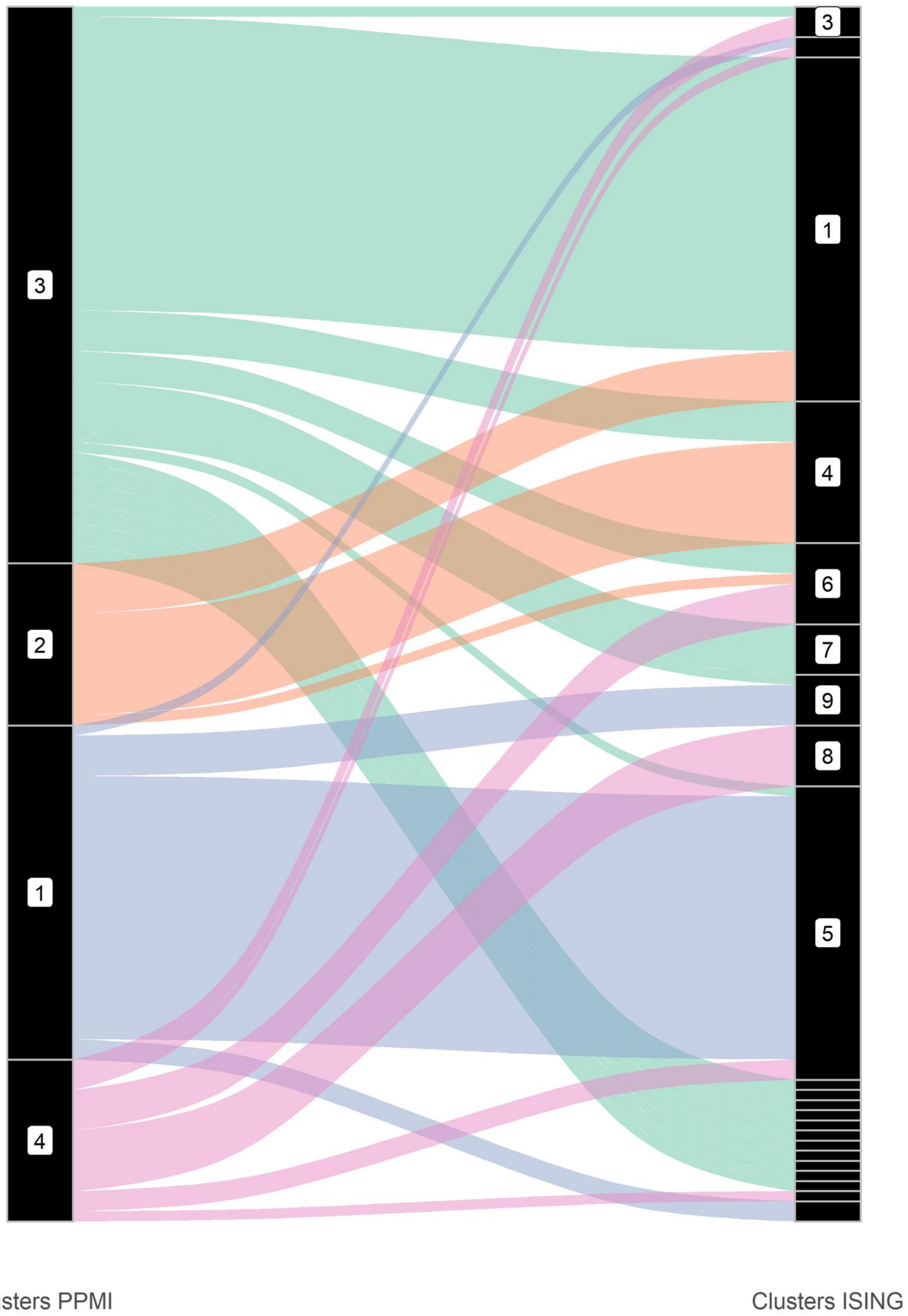


**Figure S15. Sankey showing comparison between clusters in PPMI and Ising, aripiprazole.**

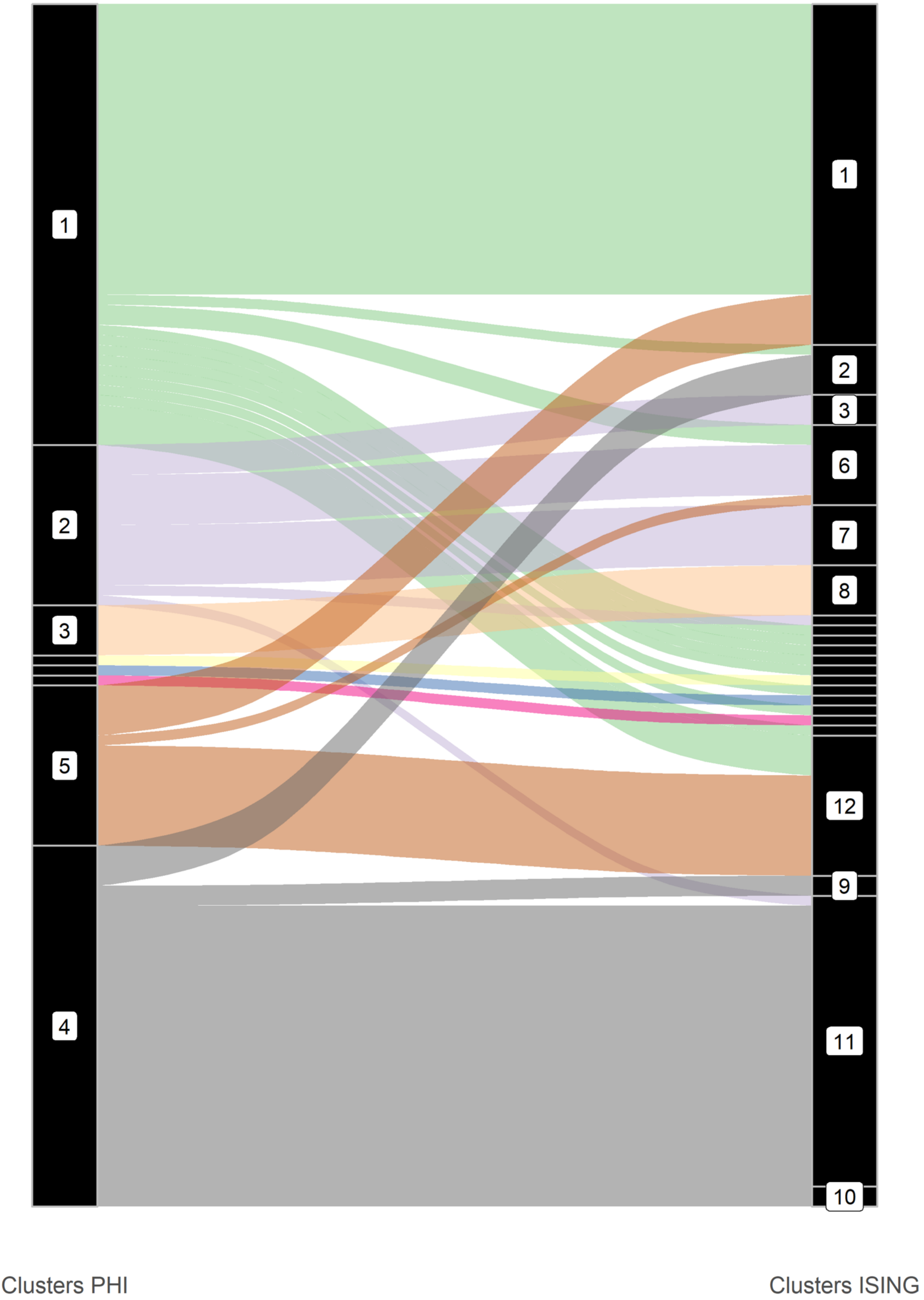


**Figure S16. Sankey showing comparison between clusters in PHI and Ising, aripiprazole.**
